# Analytical Validation of NeXT Personal^®^, an Ultra-sensitive Personalized Circulating Tumor DNA Assay

**DOI:** 10.1101/2024.01.17.23299863

**Authors:** Josette Northcott, Gabor Bartha, Jason Harris, Conan Li, Fabio C. P. Navarro, Rachel Marty Pyke, Manqing Hong, Qi Zhang, Shuyuan Ma, Tina X. Chen, Janet Lai, Nitin Udar, Juan-Sebastian Saldivar, Erin Ayash, Joshua Anderson, Jiang Li, Tiange Cui, Tu Le, Ruthie Chow, Randy Velasco, Chris Mallo, Rose Santiago, Robert C. Bruce, Laurie J. Goodman, Yi Chen, Dan Norton, Richard O. Chen, John M. Lyle

**Affiliations:** Personalis, Inc., Fremont, CA 94555, USA

## Abstract

We describe the analytical validation of NeXT Personal^®^, an ultra-sensitive, tumor-informed circulating tumor DNA (ctDNA) assay for detecting residual disease, monitoring therapy response, and detecting recurrence in patients diagnosed with solid tumor cancers. NeXT Personal uses whole genome sequencing of tumor and matched normal samples combined with advanced analytics to accurately identify up to ∼1,800 somatic variants specific to the patient’s tumor. A personalized panel is created, targeting these variants and then used to sequence cell-free DNA extracted from patient plasma samples for ultra-sensitive detection of ctDNA.

The NeXT Personal analytical validation is based on panels designed from tumor and matched normal samples from two cell lines, and from 123 patients across nine cancer types. Analytical measurements demonstrated a detection threshold of 1.67 parts per million (PPM) with a limit of detection at 95% (LOD_95_) of 3.45 PPM. NeXT Personal showed linearity over a range of 0.8 to 300,000 PPM (Pearson correlation coefficient= 0.9998). Precision varied from a coefficient of variation of 12.8% to 3.6% over a range of 25 to 25,000 PPM. The assay targets 99.9% specificity, with this validation study measuring 100% specificity and *in silico* methods giving us a confidence interval of 99.92 to 100%.

In summary, this study demonstrates NeXT Personal as an ultra-sensitive, highly quantitative and robust ctDNA assay that can be used to detect residual disease, monitor treatment response, and detect recurrence in patients.

## INTRODUCTION

Tumor DNA shed into the patient’s bloodstream can be detected as circulating tumor DNA (ctDNA). When the patient undergoes surgical resection or other treatment, ctDNA is a powerful noninvasive tool for detecting molecular residual disease (MRD), monitoring for recurrence, or tracking therapy response. For example, post-surgery, the presence of ctDNA has been shown to predict disease recurrence [1–4] and lower overall survival in a wide range of cancers [5–15]. Detection of ctDNA raises the possibility of escalating treatment for patients earlier, with the goal of improving outcomes. For example, the CIRCULATE Japan study on resectable colorectal cancer stage II-IV, looked at using ctDNA status to determine whether to escalate or de-escalate treatment post-operatively [16]. The absence of tumor post-surgery, as indicated by the absence of ctDNA, potentially allows for the de-escalation of adjuvant treatment, thereby avoiding unnecessary toxicity to the patient, as well as saving on medication costs [4, 17–20]. In the DYNAMIC trial, patients negative for ctDNA were spared adjuvant chemotherapy. The cohort containing these patients had non-inferior 3-year recurrence-free survival compared to patients who received standard adjuvant chemotherapy, suggesting that ctDNA-negative patients could forgo chemotherapy without deleterious effect [19].

Several ctDNA assays have been shown to detect tumor recurrence earlier than radiological imaging owing to their sensitivity [3, 7, 21–25]. Such ctDNA assays have also predicted metastases in seemingly non-recurrent patients. In HR+ breast cancer patients who were serially tested, positive ctDNA was detected before distant metastasis was observed [26]. Because of their specificity and sensitivity in detecting recurrent disease, ctDNA assays are also useful in clinical trial design and recruitment [22, 27]. In addition, ctDNA assays help identify patients at higher risk of recurrence, such that treatment efficacy would be enhanced, thereby reducing sample size, time to trial completion, and cost for the study. The exclusion of low ctDNA patients who are not at risk of disease progression spares those patients from exposure to inappropriate therapy. During a clinical study, ctDNA can also be an early signal of drug efficacy, though further extensive clinical validation will be required before ctDNA assays can be used as a surrogate endpoint [28].

Changes in ctDNA levels following therapy can be indicative of disease response. After chemotherapy or radiological treatment of advanced cancers, a decrease in ctDNA was correlated with better patient response in a range of cancers [20, 29–33]. Conversely, increased ctDNA was associated with shorter progression-free survival [34].

In recent years, two major approaches have emerged for detecting ctDNA in cancer patients: tumor-informed and tumor agnostic approaches. Tumor-informed approaches utilize the patient’s tumor to create a custom panel specific for detecting ctDNA based on variants found in the tumor. Tumor agnostic approaches use a fixed panel assay design for all patients, and while convenient, are generally less sensitive compared to tumor-informed assays [35]. For these current ctDNA assays, limits of detection (LODs) typically range from 0.008 to 0.25% (or 80 to 2,500 in units of parts per million, PPM) [9, 36–39]. The lower the LOD of the assay, the more sensitive it is, and the earlier a residual or recurrent tumor could be detected. For example, it has been claimed that an LOD of 0.01% (100 PPM) or lower is required to detect metastatic recurrence in triple negative breast cancer [40]. Conversely, a ctDNA assay with less sensitivity may result in undetected tumor and consequently missed treatment opportunities (41, 42]. For example, in a recent study on resected CRC patients, one ctDNA assay detected tumor recurrence 53.3% of the time, no better than imaging and carcinoembryonic antigen (CEA), which had a 60.0% detection rate. Poor sensitivity of the assay thus led the authors to conclude that the ctDNA assay may not be advantageous as a surveillance strategy for tumor recurrence compared to imaging and CEA [43].

In this paper, we present the analytical validation of NeXT Personal, a tumor-informed, ctDNA assay (Figure 1) that utilizes whole genome sequencing (WGS) and NeXT SENSE™ (Signal Enhancement and Noise Suppression Engine) to significantly improve sensitivity to ctDNA, while preserving high specificity. Starting with whole genome sequencing of a tumor/normal tissue sample from a patient, NeXT Personal designs a personalized panel of up to ∼1,800 tumor variants to enhance sensitivity compared to other tumor-informed panel approaches that typically rely on up to ∼50 variants based on whole exome sequencing (WES). This custom panel is then used to detect traces of ctDNA from a patient’s blood sample for MRD or recurrence detection.

**Figure 1.**
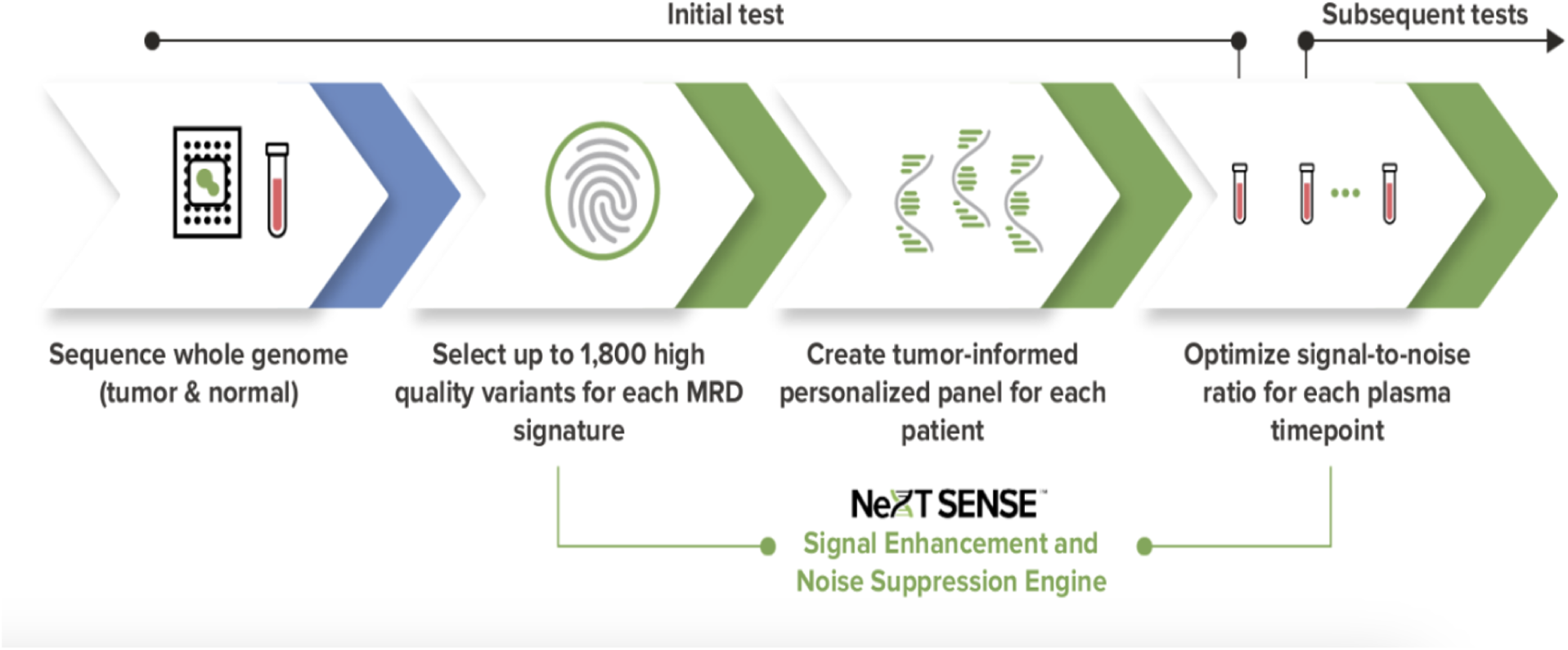
NeXT Personal tumor-informed ctDNA process

## RESULTS

### Overall performance

The performance of NeXT Personal was analytically validated by testing 123 tumor/normal/plasma sets from cancer patients, 131 donor normal plasma samples, and 2 tumor/normal matched cell line pairs, including a commercially available reference sample from SeraCare. For each sample set, a panel of up to ∼1,800 tumor-specific variants was created from WGS data from the tumor tissue and the associated normal specimen. The analytical validation consisted of the following studies: accuracy, analytical range measurements (detection threshold, limit of blank, limit of detection, precision, linearity, limit of quantification, contrived sample functional characterization), clinical sample performance, specificity, effect of interfering substances, and effect of cfDNA input amount. The overall performance characteristics of the assay are summarized in Table 1.

**Table 1.** Performance Specifications and Analytical Validation Performance of NeXT Personal.

|  |  |  |
| --- | --- | --- |
| Metric | Description | Measured Performance |
| *Assay parameters* |  |  |
| Panel size | Number of tumor-specific targets | Up to ~1,800 somatic variants |
| *Accuracy* |  |  |
| Quantitative accuracy | Agreement between measured and target value | 1.15 to 1,617 PPM<br>($r^2 = 0.9987$) |
| Sensitivity | Rate of true positive detections of known positive samples | 100% (98.5%-100%) |
| Positive predictive value (PPV) | Rate of true positive detections of total positive detections | 100% (CI 96.0%-100%) |
| Negative predictive value (NPV) | Rate of true negative detections of total negative detections | 100% (CI 98.5%-100%) |
| *Analytical range measurements* |  |  |
| Detection Threshold* | Signal threshold for making a positive call | 1.67 PPM |
| Limit of Blank* | Upper 95th percentile of signal on blank specimens | 0.719 PPM |
| Limit of Detection* | Lowest concentration detectable in 95% of replicates | 3.45 PPM |
| Precision | Coefficient of variation | 4.15% (25,000 PPM)<br>3.55% (10,000 PPM)<br>12.8% (25 PPM) |
| Linearity | Range 0.8 to 300,000 PPM | Pearson correlation coefficient = 0.9998 |
| Limit of Quantification | Lowest concentration measured with total error <25% | 10 PPM |
| *Clinical sample performance* |  |  |
| Sample processing success rate | Rate of success across 9 tumor types, stages I-IV | 99.1% |
| *Specificity* |  |  |
| Specificity | Rate of negative calls on normal samples | 100% (CI 99.92%-100%) |
| *Effect of interfering substances* |  |  |
| Genomic DNA (≥1.5 kb) | Consequence of leukocyte lysis in collected blood sample | Robust up to 25% |
| *Cell-free DNA input amount* |  |  |
| Sample input quantity | Input range of clinical samples returning results within the defined total allowable error | 2 to 30 ng |
| CI = confidence interval<br>*Representative panel, performance of individual panels may vary. |  |  |

### Accuracy

The accuracy study used an orthogonally confirmed, commercially available reference (Seraseq^®^ ctDNA MRD Panel Mix, SeraCare Life Sciences, Gaithersburg, MD) to assess the quantitative and qualitative accuracy of NeXT Personal. A personalized panel was created based on somatic variants detected in the WGS sequencing of the tumor and matched normal cell line genomic DNA (gDNA). Target variants were chosen such that the distribution of single nucleotide variant (SNV) substitutions (*e.g.*, C to T) as well as the allele frequencies mimicked those seen in typical clinical samples, and was necessary due to the atypical distribution of these features in the reference sample.

To assess the quantitative accuracy of NeXT Personal over a range of ctDNA concentrations, samples of known concentration were prepared through a serial dilution of the SeraCare reference into its matched normal. The dilution series consisted of 9 levels with ctDNA concentrations ranging from 1.15 to 1,617 PPM. At least 3 replicate samples per level were analyzed, with samples being processed by 2 operators. The ctDNA signal in each sample, as measured by NeXT Personal, is shown in Figure 2, as a function of known sample ctDNA concentration, with the identity relation shown by the solid line (y=x). Regression fit of the data yielded a correlation coefficient of 0.9987. Ordinary least squares regression was applied to the data. The 95% confidence interval (CI) for the intercept includes zero (-2.47, 3.25), and the CI for the slope includes unity (-0.99,1.01), indicating that the NeXT Personal measured results accurately reflect the known sample ctDNA concentration over the entire range of dilutions. Thus, NeXT Personal shows high analytical accuracy in measuring ctDNA concentration over that range.

**Figure 2.**
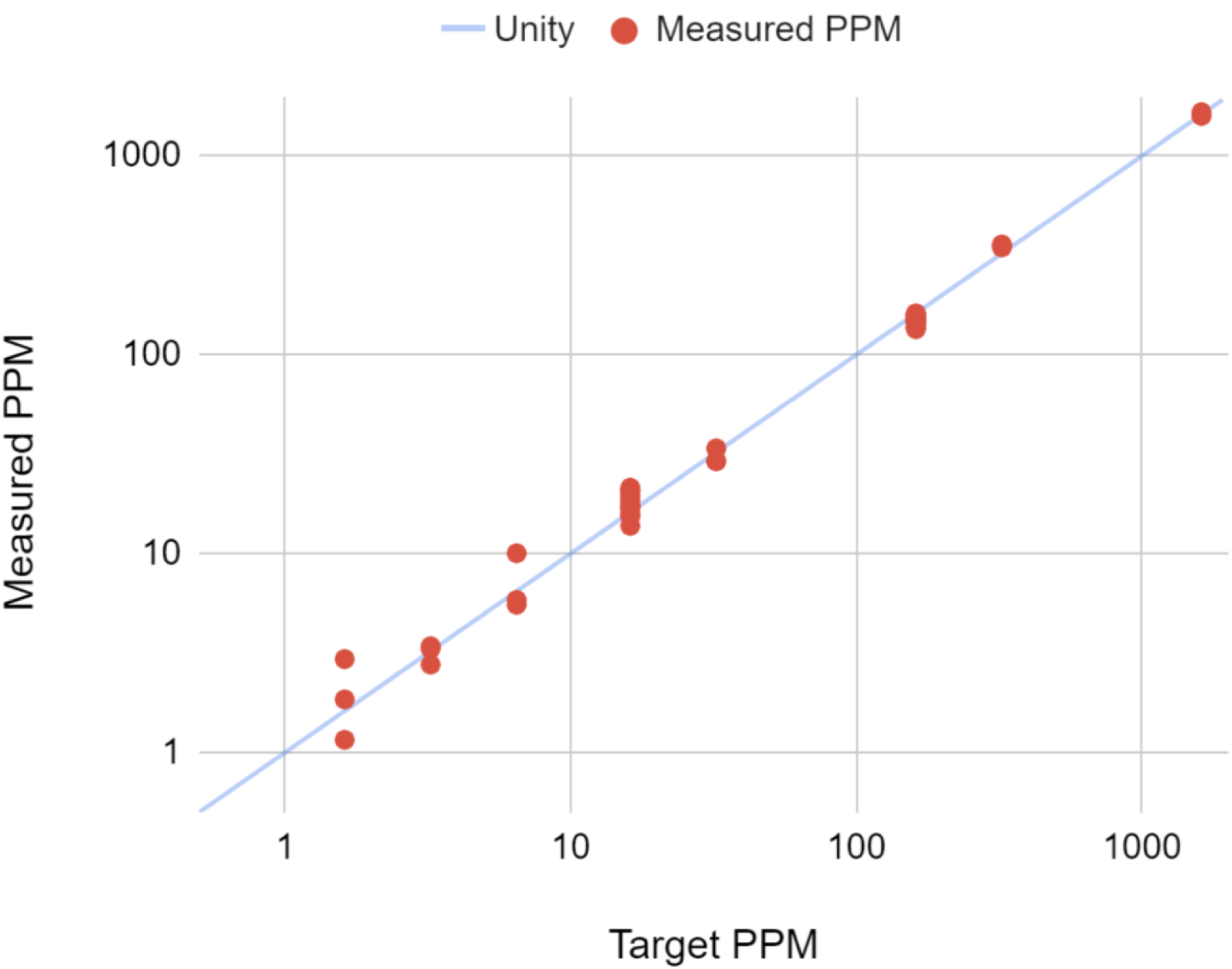
Measured ctDNA signal reported on NeXT Personal as a function of known ctDNA concentration. The solid line is the identity line (y=x).

To assess the qualitative accuracy of the assay, positive reference samples with the lowest concentrations available commercially, along with negative samples, were tested with NeXT Personal. The known positive samples consisted of 20 samples at 0.05% ctDNA and 20 samples at 0.005% ctDNA, obtained from SeraCare. Negative samples included three negative control specimens from SeraCare, 46 normal cell line cell-free DNA (cfDNA) samples (see materials and methods), as well as 239 healthy donor normal clinical specimens assayed with a NeXT Personal panel designed for an unrelated patient, for a total of 288 negative-control samples. All samples were sequenced and analyzed with the NeXT Personal assay. The analysis assigns a P-value to describe the significance of the ctDNA measurement, and if the P-value is less than 0.001, then the sample is labeled as “ctDNA Positive”; otherwise it is labeled as “ctDNA Negative”. The results are summarized in Table 2. All positive and negative control samples were correctly classified by NeXT Personal.

**Table 2.** NeXT Personal Accuracy.

| Table 2. NeXT Personal Accuracy |  |  |  |
| --- | --- | --- | --- |
|  |  | Sample Values |  |
|  |  | Positive | Negative |
| NeXT Personal | Positive | 40 | 0 |
|  | Negative | 0 | 288 |

Sensitivity, specificity, positive predictive value (PPV), and negative predictive value (NPV), were calculated as described in the supplemental materials, and the results are summarized in Table 3.

**Table 3.** NeXT Personal Performance Characteristics.

| Table 3. NeXT Personal Performance Characteristics |  |
| --- | --- |
| Analytical Sensitivity (95% CI) | 100% (98.5%-100%) |
| Analytical Specificity (95% CI) | 100% (96.0%-100%) |
| Positive Predictive Value (95% CI) | 100% (96.0%-100%) |
| Negative Predictive Value (95% CI) | 100% (98.5%-100%) |

### Analytical Range Measurements

NeXT Personal is a quantitative assay, providing a readout of the ctDNA signal of a plasma sample, in PPM. The analytical range of the assay helps users understand when multiple ctDNA measurements can be considered to be quantitatively different from each other, enabling quantitative monitoring of treatment response or disease burden.

To demonstrate the analytical range of NeXT Personal we used a contrived sample set created from cfDNA derived from a breast tumor cell line and a patient-matched normal cell line (see materials and methods). We created a NeXT Personal panel for these studies from WGS of the same tumor and normal cell line gDNA. The panel variants were verified to ensure that the SNV substitutions and allele frequencies closely mimicked a typical clinical sample’s panel. This NeXT Personal panel was used on the contrived sample set, as well as on a set of healthy donor normal samples, to measure the detection threshold, limit of blank (LOB), 95th percentile limit of detection (LOD_95_), limit of quantification (LOQ), precision, and linearity of the assay (see Discussion). While the detection threshold, LOB and LOD_95_ values are those of a representative panel and dependent on the panel used, the precision, linearity and LOQ can be considered as defining the performance of the assay itself, and are not dependent on individual panel designs.

### Detection Threshold

For any given measurement, the detection threshold of NeXT Personal varies according to the parameters of the panel as well as the amount of cfDNA input into the assay. As stated above, measurements are called positive for ctDNA detection when the probability of the call having resulted from noise is <0.001, thus meeting our specificity requirement of >99.9%. As part of the NeXT Personal readout, we compute the ctDNA signal at the detection threshold. For the 190 measurements performed in the analytical range measurement studies, the detection threshold ranged from 1.47 to 1.87 PPM (1.67 PPM mean).

### Limit of Blank

The limit of blank (LOB) for a ctDNA assay is the ctDNA noise signal measured in donor normal samples. For a tumor-informed assay such as NeXT Personal, this is appropriately characterized both for an individual panel, as described in this study, or for a set of panels, such as was done for the Specificity study described below. Characterization of the LOB for the contrived sample panel allows us to use the LOB measurement to derive the LOD_95_ in the subsequent study.

The limit of blank was measured using a collection of cfDNA from 121 donor normal plasmas and 46 normal cell line cfDNA samples (see materials and methods). The samples in this study were processed on different days, by different operators and using two reagent lots of the hybridization-capture enrichment kit (Twist Bioscience, South San Francisco, CA). A Shapiro-Wilk test demonstrated significant departure of the data from normality and hence a non-parametric approach was taken. The results for two hybridization-capture enrichment kit lots are shown. The higher value of the two results, 0.719 PPM, is taken as the LOB [44].

### Limit of Detection

The limit of detection (LOD_95_) is defined as the ctDNA concentration at which 95% of measurements will yield a positive result [44]. In the LOD_95_ study, the contrived sample system was used to create a series of 7 low positive cfDNA samples with known ctDNA concentrations ranging from 0.8 to 8.2 PPM. Each of the 7 cfDNA samples was analyzed with NeXT Personal 5 times by 2 operators, for a total of 70 runs. To capture assay variability, the 5 replicate cfDNA libraries for each operator were enriched by the operator over 2 days, using different reagent lots of the hybridization-capture kit (Twist Bioscience) on each of the days. Using the precision profile approach (see Supplementary Materials) [44], the LOD_95_ was calculated for each reagent lot (Table 5). The final LOD_95_ determination is 3.45 PPM and is based on selecting the highest of the reagent lot estimates. Note that while this represents the lowest concentration at which 95% of positive samples are detected, any positive call above the detection threshold (∼1.67 PPM for this panel) meets our specificity requirement, having less than 1 in 1000 probability of being a false positive.

**Table 4.** Limit of blank (LOB) on two reagent lots.

| Table 4. Limit of blank (LOB) on two reagent lots. |  |  |
| --- | --- | --- |
|  | Enrichment Reagents Lot A | Enrichment Reagents Lot B |
| Number of samples | 82 | 85 |
| Upper 95<sup>th</sup> percentile ctDNA signal of blank samples | 0.493 PPM | 0.719 PPM* |
| *The higher value of the two reagent lot results is taken as the limit of blank. |  |  |

**Table 5.** Calculated LOD for Each Reagent Lot.

| Table 5. Calculated LOD for Each Reagent Lot |  |  |
| --- | --- | --- |
|  | Enrichment Reagents Lot A | Enrichment Reagents Lot B |
| LOD (PPM) | 1.80 | 3.45 |

### Precision

The goal of this study was to characterize the variability of the NeXT Personal ctDNA measurements among replicate samples at a range of potential signal levels. The total variance observed provides an estimate of how different two signals from a set of patient samples need be before they can be considered analytically different. Levels of 25, 1,000, and 25,000 PPM were selected as representative of low, medium, and high ctDNA signal levels in clinical samples (supplemental Figure S1). Each of the three contrived cfDNA samples containing ctDNA at the selected signal levels was analyzed with NeXT Personal 24 times. To capture the most potential assay variability, the cfDNA samples were quantified by 2 operators, each using two different Cell-free DNA ScreenTape Analysis (Agilent Technologies) reagent lots prior to library preparation, and the pre-enrichment cfDNA libraries were enriched by 2 operators over 3 days using two different reagent lots of the hybridization-capture enrichment kit (Twist Bioscience). The overall precision was calculated as the coefficient of variation (CV), or the standard deviation divided by the mean value, over all runs for a given ctDNA concentration level.

A multivariate analysis of variance applied to the results did not point to any single factor as an overriding contributor to variability. The highest CV was observed at the lowest signal level (Table 6). The 95% confidence interval is 1.956 standard deviations of the measurement value, which at 25 PPM is 24.98%. Thus, we are using 25% as the total allowable error (TAE) for measurements in this study.

**Table 6.** Precision Summary.

| Table 6. Precision Summary |  |  |  |  |
| --- | --- | --- | --- | --- |
| Sample | Total runs | Mean concentration, measured (PPM) | Standard deviation (PPM) | Coefficient of variation (%) |
| High concentration (~25,000 PPM) | 24 | 25,017 | 1,038.2 | 4.15% |
| Medium concentration (~1,000 PPM) | 24 | 991.0 | 35.18 | 3.55% |
| Low concentration (~25 PPM) | 24 | 26.02 | 3.323 | 12.77% |

### Linearity

The purpose of this study was to assess the linearity of the NeXT Personal assay. In order to determine how well the measured ctDNA signals correlate with the known ctDNA concentrations, a series of diluted samples were prepared at 19 different ctDNA concentrations ranging from 0.8 PPM to ∼300,000 PPM. A minimum of 3 replicate samples at each ctDNA concentration level were tested with NeXT Personal, and the measured ctDNA signals are plotted against the known ctDNA concentration on a log-log scale, in

### Limit of Quantification

The limit of quantification (LOQ) is the lowest amount of a ctDNA signal that can be detected in a sample at a given level of accuracy. In this assay, we define the LOQ as the point at which a difference greater than 25% (the TAE) could be observed due to variability between two observations of the same ctDNA concentration. To determine the LOQ, 6 cfDNA samples were prepared using the contrived sample system to attain ctDNA concentrations of 6.6, 8.2, 10, 17, 25 and 50 PPM. Each of the 6 cfDNA samples was assayed 10 times with NeXT Personal. The 10 replicate cfDNA libraries at each ctDNA concentration were enriched by 2 operators over 2 days using different reagent lots of the hybridization-capture kit (Twist Bioscience) on each of the days. The mean of the measured ctDNA signal, in PPM, was compared to the target concentration, and the bias and standard deviations of the replicate measurements were also determined. The results are shown in Table 7. The lowest ctDNA concentration for which the total error is <25% for both reagent lots is chosen for the final LOQ, thus, the final LOQ was determined to be 10 PPM.

**Table 7.** Analysis of LOQ results by reagent lot (A,B)

| Table 7. Analysis of LOQ results by reagent lot (A,B) |  |  |  |  |
| --- | --- | --- | --- | --- |
| Target PPM | Bias between measured and target PPM | Standard deviation of measured values (PPM) | Total Error (TE) in PPM (root mean square) | TE, % |
| 50 | 0.87, -2 | 3.77, 3.78 | 3.86, 4.27 | 7.73%, 8.55% |
| 25 | 0.64, 1.35 | 3.05, 2.83 | 3.11, 3.13 | 12.5%, 12.5% |
| 17 | 1.78, -0.31 | 2.13, 2.35 | 2.78, 2.37 | 16.4%, 13.9% |
| **10** | 0.74, 0.26 | 1.1, 1.73 | 1.33, 1.75 | 13.3%, 17.5% |
| 8.2 | 1.13, -0.23 | 2.07, 0.99 | 2.36, 1.02 | **28.8%\***, 12.5% |
| 6.6 | -0.13, 1.56 | 1.93, 0.88 | 1.93, 1.79 | **29.3%, 27.1%\*** |
| *The limit of quantification is set as the lowest ctDNA concentration for which total error is <25% for both reagent lots. |  |  |  |  |

### Contrived Sample Functional Characterization

To demonstrate that the contrived cell line cfDNA samples used for the analytical range studies perform similarly to clinical samples, we conducted the contrived sample functional characterization study. In addition to the contrived samples described above, two sets of clinical cfDNA samples were prepared by serial dilution of ctDNA positive patient cfDNA with donor normal cfDNA to achieve samples with ctDNA concentrations ranging from 1.2 to 25 PPM. The clinical cfDNA samples originated from a breast cancer patient (Cureline, Brisbane, CA) and a non-small cell lung cancer patient (iProcess, Irving, TX).

The contrived samples at each ctDNA concentration were assayed 10 times or more with NeXT Personal and the average ctDNA signal, in PPM, was plotted against the expected ctDNA concentration on the same x-y graph as the clinical samples, which were assayed a single time at each ctDNA concentration point (Figure 4).

**Figure 3.**
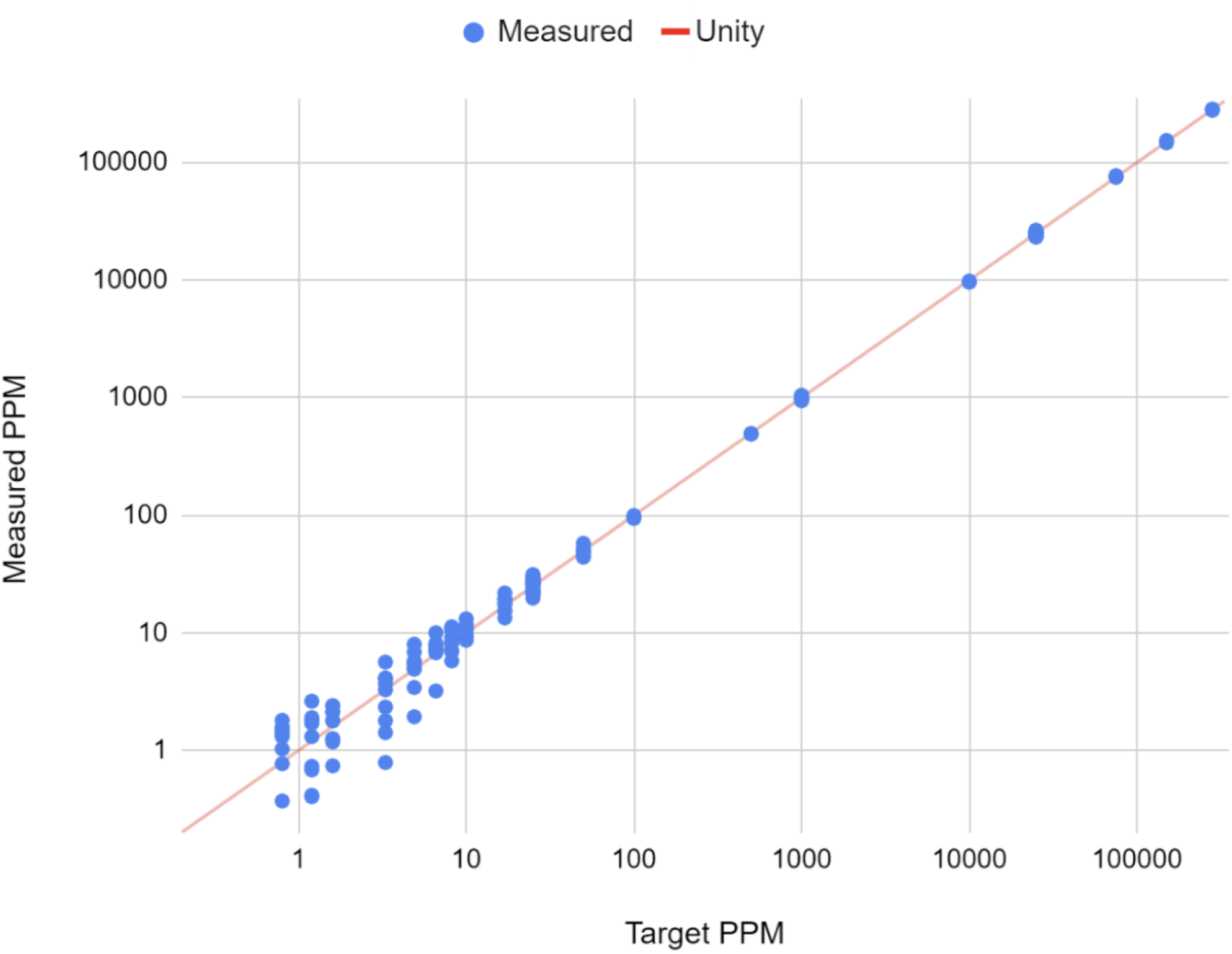
The line shown is the identity line (y=x). A regression fit of the data yielded a Pearson correlation coefficient of 0.9998 (p<0.001). This indicates that measured and input concentrations were linearly correlated over the range of 0.8 to 300,000 PPM. Measured ctDNA signal versus target ctDNA concentration (PPM), showing the linearity of the NeXT Personal assay.

**Figure 4.**
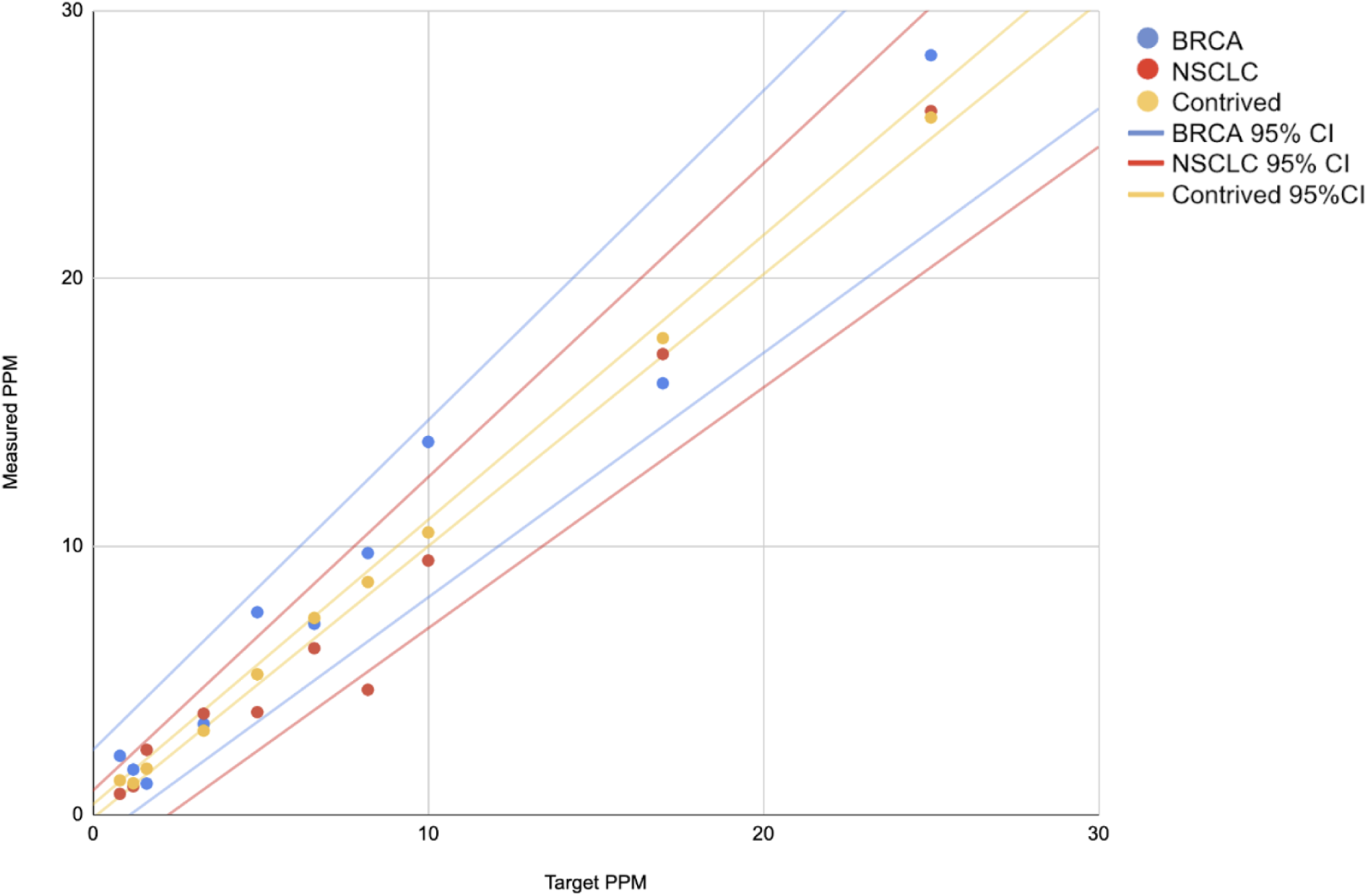
The plot of measured ctDNA concentration versus expected (target) concentration is shown for the contrived cell line and two clinical samples. The 95th percentile confidence intervals for the samples are shown as matching colored lines.

To demonstrate equivalence, the slopes and intercepts of the regression fits were compared between the cell line samples and clinical samples. The results are summarized in Table 8. There is no significant difference between the contrived sample values and clinical sample values, with the 95% confidence intervals of the slopes and intercepts for both clinical samples fully encompassing the slope and intercept for the contrived sample. Based on the overlap in regression intercept and slope values of the contrived cell line and clinical samples, we conclude that the two types of samples are functionally comparable, and therefore the use of cell line samples in the validation study is appropriate.

**Table 8.** Linear regression slopes and intercepts from X-Y plots of measured concentration versus expected concentration.

|  |  |  |
| --- | --- | --- |
| Sample | Linear regression intercept value<br>(95% CI), PPM | Linear regression slope<br>value (95% CI) |
| Contrived sample | 0.120 (-0.14, 0.37) | 1.04 (1.01, 1.06) |
| Breast Cancer (BC) Clinical sample | 0.69 (-1.02, 2.41) | 1.07 (0.91, 1.23) |
| Lung Cancer (LC) Clinical sample | -0.57 (-2.03, 0.89) | 1.03 (0.90, 1.17) |

### Clinical Sample Performance

The purpose of this study was to demonstrate the end-to-end performance of NeXT Personal on clinical samples. This study was conducted using clinical specimen sets for which tumor tissue, normal tissue, and plasma samples, collected at a pre-surgical time point, were available (various vendors, see materials and methods). A total of 118 clinical sample sets were assayed, representing nine different cancer types (Table 9 and Figure 5). Two specimen sets were found to be from the same patient, resulting in 117 unique patients in the dataset (Table A).

**Figure 5A:**
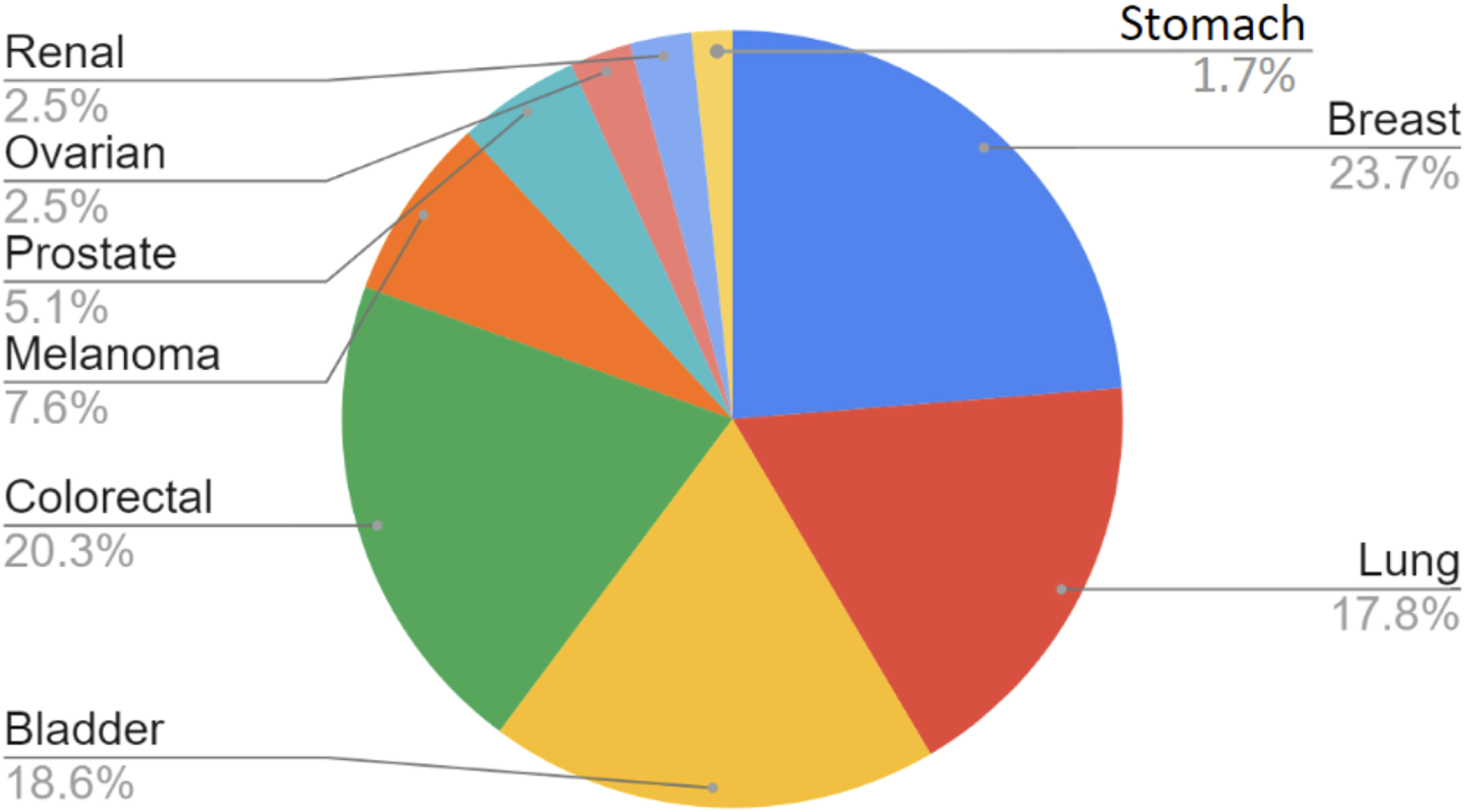
Cancer type of tumor samples used in the clinical sample performance study

**Figure 5B:**
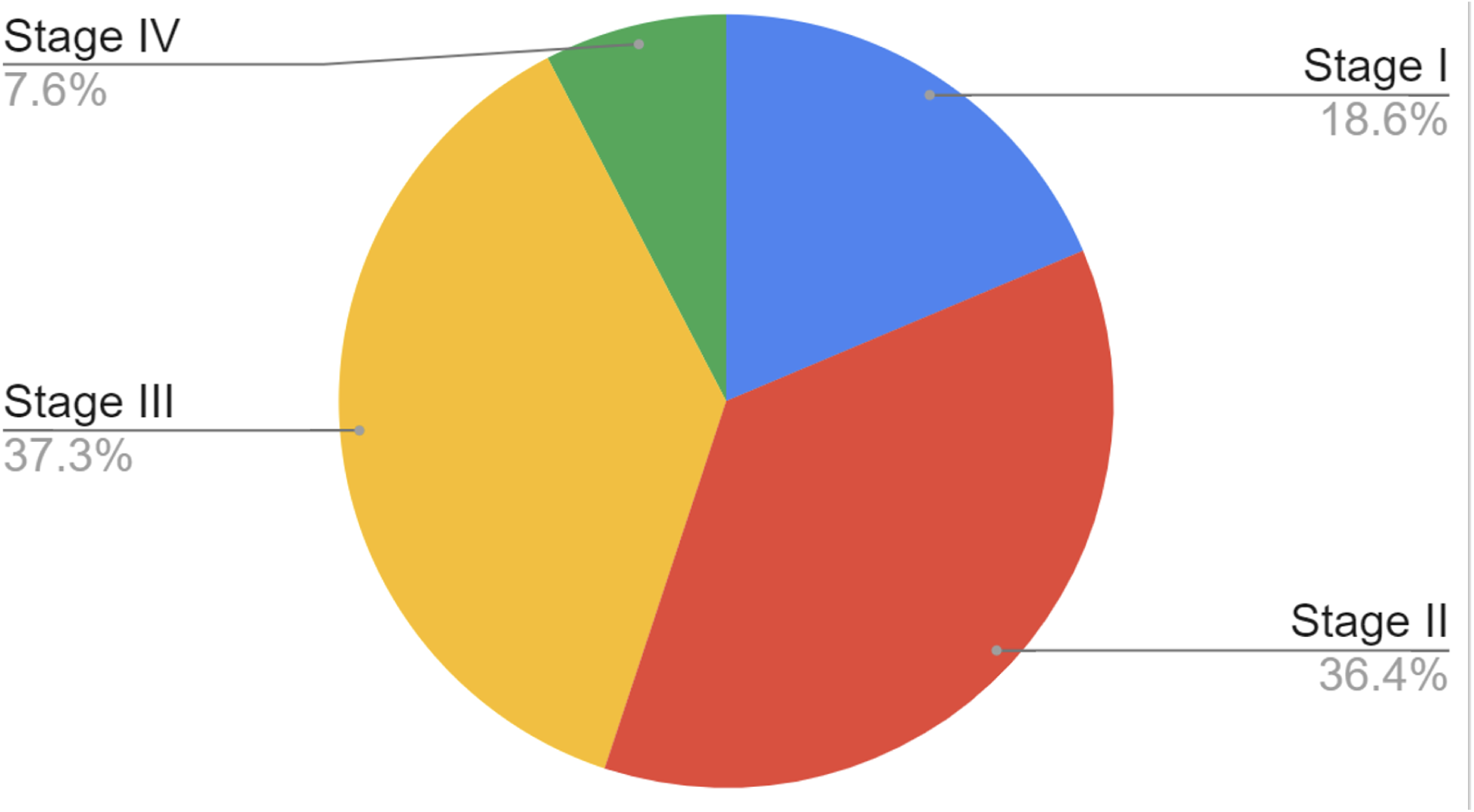
Cancer stage of tumor samples used in the clinical sample performance study.

**Table 9:** Clinical samples used in the NeXT Personal clinical sample performance study.

|  |  |  |  |  |
| --- | --- | --- | --- | --- |
|  | Stage |  |  |  |
| Type | I | II | III | IV |
| Lung | 4 | 10 | 6 | 1 |
| Breast | 9 | 12 | 6 | 1 |
| Bladder | 3 | 7 | 8 | 4 |
| Colorectal | 5* | 7 | 12 | 0 |
| Melanoma | 1 | 3 | 5 | 0 |
| Prostate | 0 | 3 | 1 | 2 |
| Ovarian | 0 | 1 | 2 | 0 |
| Renal | 0 | 0 | 2 | 1 |
| Stomach | 0 | 0 | 2 | 0 |
| *Two specimen sets were found to be from the same patient. |  |  |  |  |

Assay variability was captured by using two different reagent lots for the Cell-free DNA ScreenTape Analysis (Agilent Technologies) for cfDNA quantification prior to library preparation, and two different reagent lots of the hybridization-capture enrichment kit (Twist Bioscience) for cfDNA library enrichment with the tumor-informed panels. Furthermore, both of these sample processing steps were performed by 2 operators over multiple days. All 118 tumor samples had ≥20% tumor content, as determined by pathologists. Macrodissection of the FFPE sections was performed prior to genomic DNA extraction when it could increase the tumor content of the resulting sample. All tumor samples yielded sufficient DNA to move forward with NeXT Personal panel design. Nearly half of the 118 patient-specific panels had >1,800 somatic variant targets, with 70% of the panels having >1,000 somatic variant targets and over 85% having >500 somatic variant targets. These panels had a median detection threshold of 2.0 PPM (range 1.0 - 12 PPM). Extraction of cfDNA from up to 4.1 mL (minimum 2.1, median 3.5 mL) of plasma produced sufficient cfDNA for analysis (range 3.4 to 420 ng). Of the 118 clinical specimen sets, 5 had plasma that did not match the provided tumor/normal tissue samples. As all evidence suggested a vendor mistake, these 5 cases were excluded from the study. One plasma sample was failed due to detection of a contaminant in the sample. Therefore, in this study, the processing success rate was 99.1% (112 samples out of 113 were successfully processed).

### Specificity

In the specificity study, normal plasma samples (“blank” samples) were assayed with the NeXT Personal panels described in the clinical sample performance study to detect the presence of patient-specific tumor variants. Normal blood samples were collected in Streck cfDNA tubes from 118 donors at the Stanford Blood Center (Stanford, CA) and processed to plasma for use in these studies. Figure 6 describes the characteristics of the normal blood donors.

**Figure 6A:**
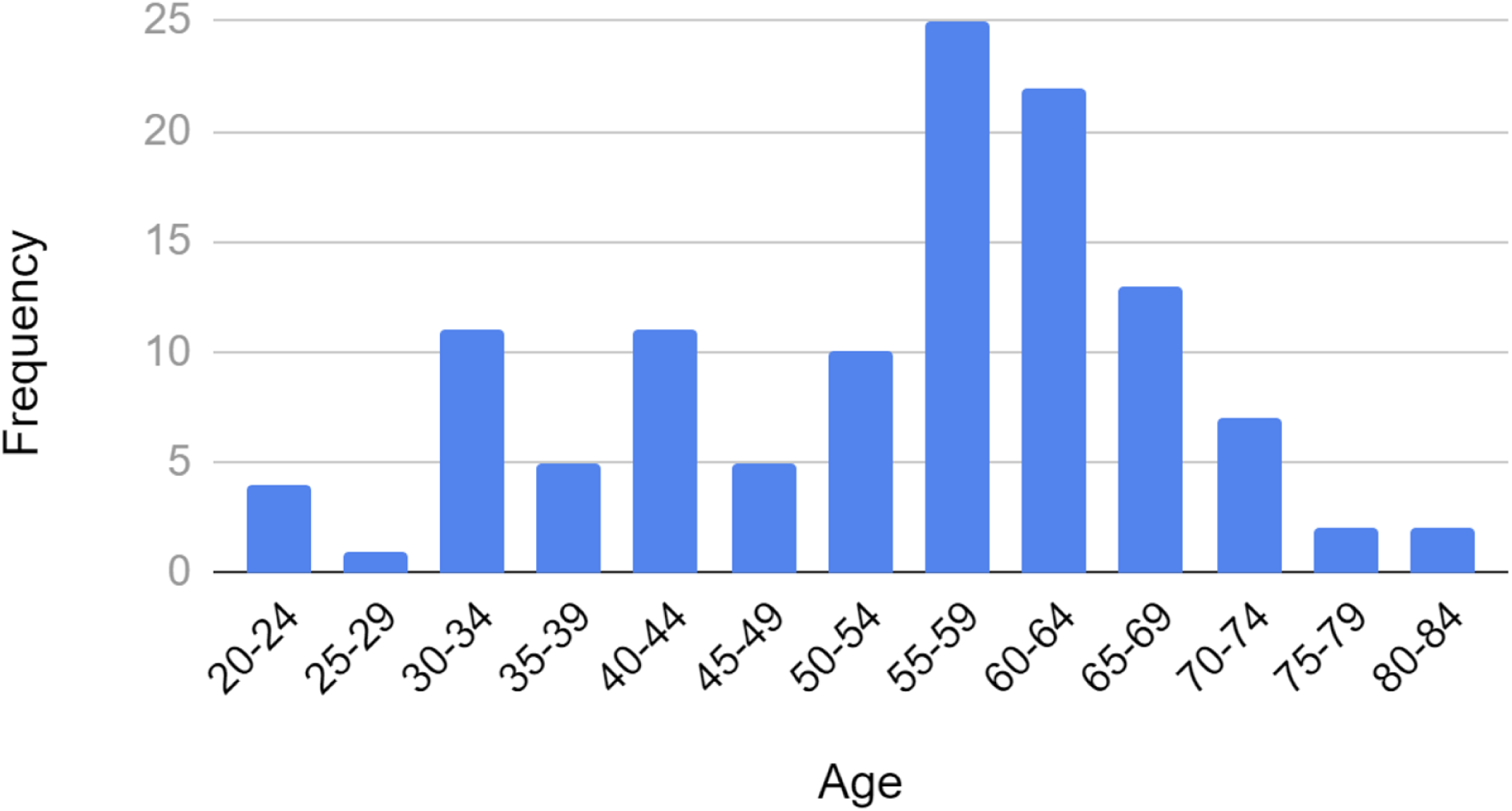
Age distribution of donor normal samples used in specificity studies.

**Figure 6B:**
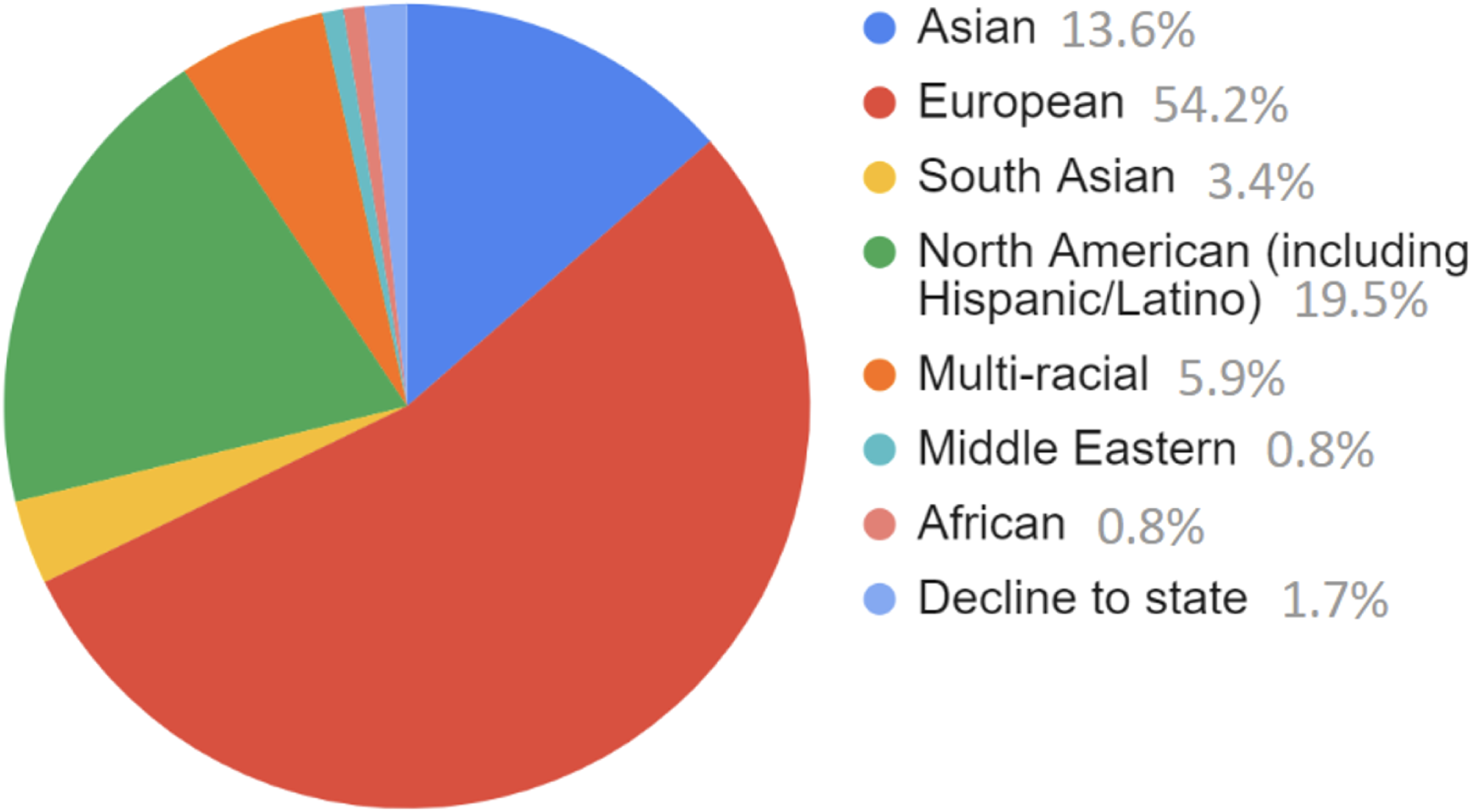
Self-reported demographics of donor normal samples used in specificity studies.1.

Per industry convention [44], sample processing for this study was performed by 2 operators over multiple days using two different reagent lots for cfDNA quantification and library enrichment. Specificity was 100%, as measured by this study (95% CI 96.8 to 100%).

Two *in silico* approaches were used to more precisely describe the specificity of the assay, the method of Abbosh *et al* (2023) [45] that draws on loci flanking each targeted site and an internally developed method relying on somatic variant target reshuffling (see materials and methods). These analyses each resulted in 23,600 *in silico* panels with the detection of 4 and 31 false positives, respectively, giving measured specificities of 99.98% (95% CI 99.96% to 100.00%) and 99.95% (95% CI 99.92% to 99.98%).

### Interfering Substances

#### Genomic DNA

The most common contaminant that could interfere with the NeXT Personal assay is genomic DNA (gDNA) from leukocyte lysis, which can occur at any point between blood collection and cfDNA extraction. While gDNA is usually observed to be quite high in molecular weight (>10 kb), in this study we chose to look at a contaminating gDNA species around 1.5 kb, which represented the shortest gDNA species observed in over a hundred patient cfDNAs extracted from plasma following Personalis standard operating procedures. The contaminating gDNA in this study was produced from high molecular weight gDNA from NA12878 (Coreill, Camden, NJ). Contrived cell line cfDNA samples containing ctDNA levels of 25, 1,000, and 25,000 PPM were spiked with 25% gDNA, or used without contaminating gDNA as control. Note that 25% represented the highest level of contamination observed in the aforementioned patient plasma samples. Samples were tested on NeXT Personal in replicates of 5. The mean concentration differences between spiked and unspiked samples are summarized in Table 11.

**Table 10:** Specificity Results.

| Table 10: Specificity Results |  |  |
| --- | --- | --- |
| Approach | Panels Tested | Specificity (95% CI) |
| Empirical measure with donor normals | 118 | 100% (96.8%, 100%) |
| *In silico* flanking | 23,600 | 99.98% (99.96%, 100%) |
| *In silico* reshuffling | 23,600 | 99.95% (99.92%, 99.98%) |

**Table 11.** Effect of gDNA spiking on ctDNA signal measurements.

| Table 11. Effect of gDNA spiking on ctDNA signal measurements |  |
| --- | --- |
| Concentration Level | Mean difference between gDNA-spiked and unspiked groups (%) |
| High concentration (25,000 PPM) | -9.33% |
| Medium concentration (1,000 PPM) | -4.29% |
| Low concentration (25 PPM) | -8.47% |

These mean differences fall below the variance of 12.77% observed in the precision study, suggesting a limited impact on determinations of ctDNA concentration due to the presence of gDNA in the sample.

In addition to gDNA contamination, we investigated the effects of hemoglobin contamination and the carryover of the final wash buffer used in cfDNA extraction. Neither of these showed significant effects (see supplemental materials).

### Effect of cfDNA input amount on ctDNA measurements

We examined the impact of cfDNA input amount on the measured ctDNA signal, in PPM. In these studies we titrated the cfDNA input from 2 to 30 ng. The modal cfDNA input for the NeXT Personal assay is approximately 15 ng cfDNA and therefore this amount was chosen as the input level for comparison. Clinical cfDNA samples from patients with breast cancer, colorectal cancer, non-small cell lung cancer (NSCLC), melanoma, and renal cancer were diluted to a ctDNA concentration of approximately 25-30 PPM and tested in triplicate with NeXT Personal. The contrived sample system was used to examine cfDNA input across the same range at 25, 1000, and 25,000 PPM measured 5 times at each point. Figure 7 and supplemental Table S1 show the NeXT Personal results from this study.

**Figure 7.**
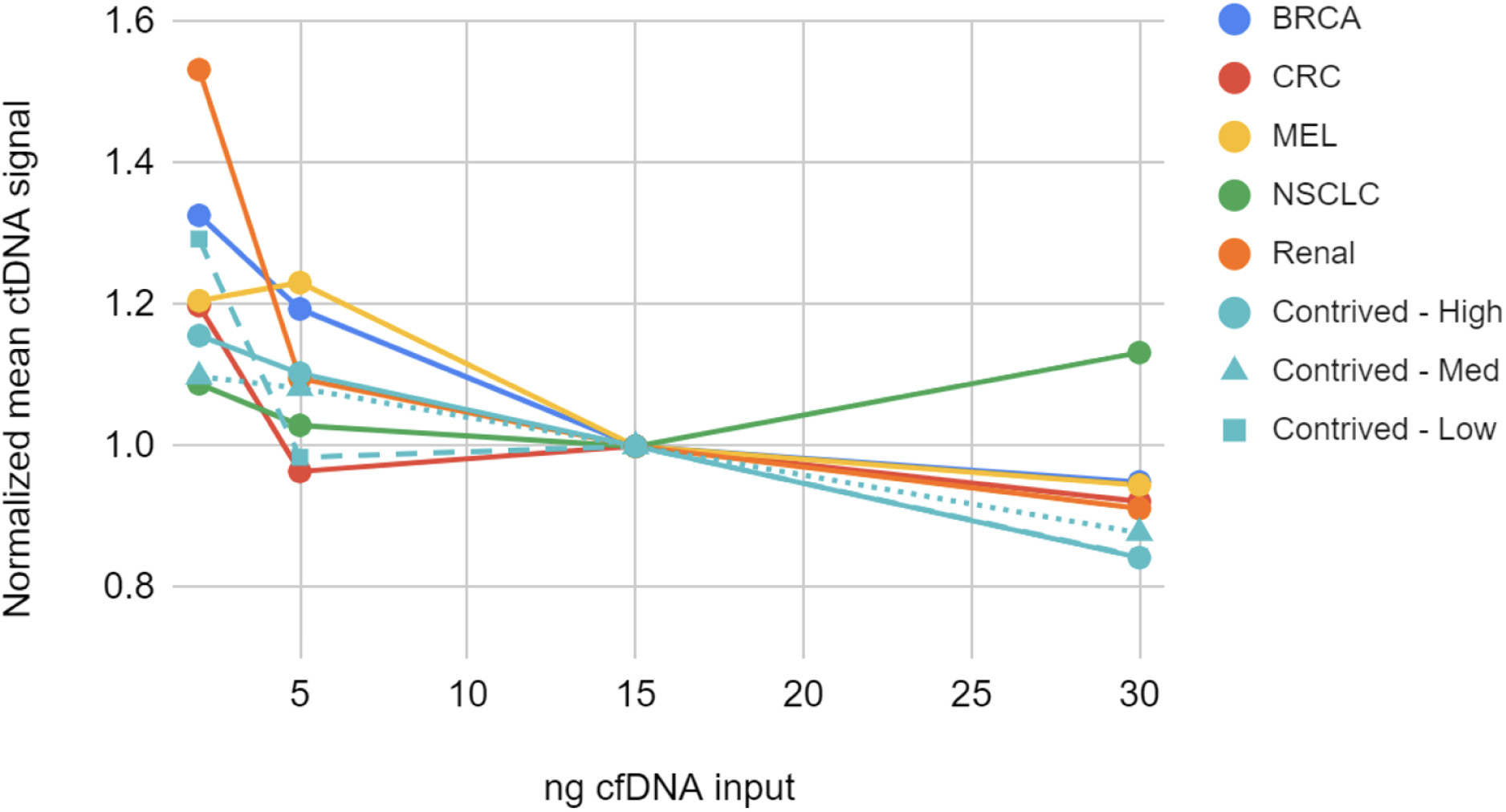
Normalized mean ctDNA signal for 5 cancer samples and 3 contrived samples at various cfDNA input quantities. Points represent the mean of replicates normalized to the 15ng input level. We observed that the measured ctDNA signal trends upward as the cfDNA input amount decreases. For the contrived samples, only the 2 ng cfDNA input samples showed significant deviation from the standard 15 ng input samples. Similarly, for the clinical samples, the 2 ng input differed significantly from the 15 ng input for the breast cancer, melanoma, and renal cancer samples. The deviation of all clinical sample points in the 2 to 30 ng range fell within the TAE (see supplemental materials). From this study we conclude that the measured ctDNA signal is not significantly affected over an input range of 5 to 30 ng.

## DISCUSSION

Effective clinical detection of ctDNA is dependent upon a sensitive and highly specific quantitative diagnostic assay with robust analytical performance. We demonstrate the analytical performance of NeXT Personal in a comprehensive manner, characterizing the limit of blank (LOB), limit of detection (LOD_95_), limit of quantification (LOQ), precision, and linearity, as well as demonstrating the specificity of the assay on a large set of clinical samples.

A commercially available control along with a large set of clinical normal samples were used to establish the overall quantitative and qualitative accuracy of the assay. The ctDNA measurements showed excellent agreement to known ctDNA concentrations over a range of 1.15 to 1,617 PPM (regression correlation coefficient of 0.9987) and the sensitivity, specificity, PPV, and NPV were measured to be 100%. Two *in silico* approaches to precisely define specificity agreed well with our targeted specificity of 99.9%, which is enabled by our proprietary noise reduction algorithms.

A contrived sample system was used to characterize the analytical range of the assay (Figure 8), demonstrating the capability of the assay to make quantitative determinations of differences in the concentrations of ctDNA present. An overview of the analytical range results are shown in Figure 8. The LOB was determined to be 0.719 PPM, established by processing 167 blank samples. The detection threshold at 99.9% specificity for these studies (1.67 PPM) was similar to that observed in the clinical sample set (median 2.0 PPM). The LOD_95_ is 3.45 PPM. This high level of sensitivity holds the promise of earlier detection of residual or recurrent disease and potentially better outcomes for the patient.

**Figure 8.**
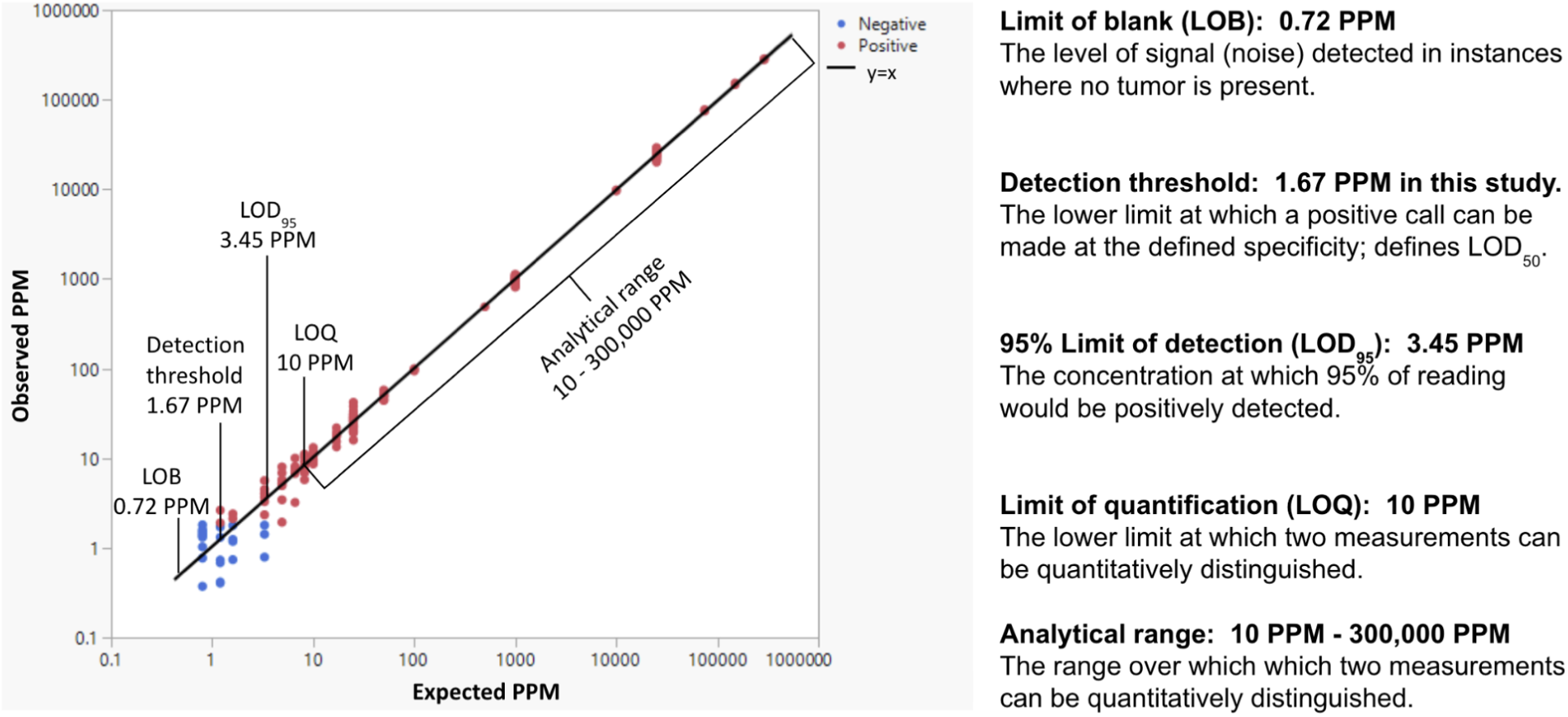
Major analytical performance measurements of the NeXT Personal assay.

Importantly, the detection of ctDNA by NeXT Personal has been shown to be linear over a wide range, from 0.8 to 300,000 PPM. Assay precision ranged from a coefficient of variation (CV) of 3.6% to 12.8%, with the highest CV at the lowest ctDNA concentration. We established a total allowable error (TAE) of 25% based on our precision measurements. We then determined the LOQ, the “lowest amount of a measurand in a material that can be quantitatively determined with stated accuracy [*i.e., the total allowable error (TAE)*]” [44]. This resulted in an LOQ of 10 PPM. This robust determination of LOQ allows calls above 10 PPM, differing by more than 25%, to be considered analytically different.

The assay was shown to perform well with clinical samples, across a range of tumor types and stages, as well as cfDNA input amounts, with a sample processing success rate of 99.1%. Further, the assay retains its quantitative capabilities across the anticipated range of cfDNA inputs.

NeXT Personal’s tumor-informed ctDNA detection approach, combining whole genome sequencing with NeXT SENSE, results in an ultra-sensitive and highly specific quantitative diagnostic assay. We have demonstrated the analytical performance of NeXT Personal across all critical aspects of a ctDNA assay. The high level of sensitivity and specificity of NeXT Personal has the potential to improve both lead times to recurrence detection and detection of molecular residual disease. The quantitative aspect of the assay is a differentiating feature which can enable more robust disease and treatment monitoring. These results suggest strong potential for clinical use of the assay in ctDNA monitoring of solid tumor cancers.

## MATERIALS AND METHODS

The accuracy, specificity, analytical range measurements and clinical sample performance studies were conducted in the Clinical Laboratory Improvement Amendments (CLIA)-certified and College of American Pathologists (CAP)-accredited laboratories at Personalis, Inc., as guided by the Association for Molecular Pathology (AMP) and CAP’s joint recommendations and following the standard operating procedures at Personalis. Interfering substances and cfDNA input amount experiments were conducted in the Personalis R&D laboratories and followed similar procedures and practices.

### Sample acquisition

Healthy donor plasma was obtained from the Stanford Blood Center (Stanford, CA). Matched sets of patient FFPE tumor tissue, buffy coat or adjacent normal FFPE tissue, and plasma were sourced from BOCA Biolistics (Pompano Beach, FL), BioOptions (Brea, CA), Cureline (Brisbane, CA), Discovery Life Sciences (Huntsville, AL), DxBioSamples (San Diego, CA), and Us4Cure (Agoura Hills, CA). Cell lines were purchased from the American Type Culture Collection and maintained under the recommended conditions. The commercially available MRD control (Seraseq ctDNA MRD Panel Mix, LGC SeraCare) was sourced directly from LGC SeraCare (Milford, MA).

### NeXT Personal Probe Panel Design

NeXT Personal custom probe panels were designed from WGS of matched tumor and normal samples using the NeXT Personal platform. Purchased FFPE tissues and buffy coat samples were processed in the Personalis CLIA/CAP laboratories following the standard operating procedures at Personalis. Briefly, one hematoxylin and eosin (H&E) stained slide was prepared from each FFPE block and used to determine the tumor content of the tissue sample. For tumor samples, macrodissection was performed when it could increase the tumor content of the resulting sample. Genomic DNA was extracted from the tissue sections and the buffy coat samples using commercially available kits (QIAGEN, Germantown, MD). DNA libraries were prepared with 50 ng to 500 ng of acoustically sheared genomic DNA (Covaris LLC, Woburn, MA) using a commercially available KAPA Kit (Roche Sequencing Solutions, Pleasanton, CA) and Personalis-optimized workflows. Whole genome sequencing was performed using NovaSeq instruments (Illumina, San Diego, California) to a depth of at least 30x. Germline and CHIP exclusion, somatic variant calling, ctDNA target selection (up to 1,800 high-quality and low-noise somatic variants), and probe design were completed using the proprietary algorithms of the NeXT Personal platform.

### Preparation of contrived cell line cfDNA samples

The HCC1954 (CRL-2338, stage IIA, invasive breast ductal carcinoma) and HCC1954BL (CRL-2339, B lymphoblast) patient-matched cell lines were maintained in RPMI 1640 media with 10% Fetal Bovine Serum. These cells were incubated in a proprietary media system prior to the collection of cfDNA from the media. Extraction of cfDNA from the cell culture media was performed using a commercially available kit (QIAGEN, Germantown, MD) and size selected with AMPure XP beads (Beckman Coulter, Indianapolis, IN) to recapitulate the cfDNA fragment sizes observed in patient plasma samples. Quantification of the cfDNA was performed with the Qubit dsDNA BR assay (Thermo Fisher Scientific, Fremont, CA) and Cell-free DNA ScreenTape Analysis (Agilent Technologies, Santa Clara, CA). Subsequently, cfDNA derived from HCC1954 was serially diluted with cfDNA derived from HCC1954-BL. Except as noted, all pre-enrichment libraries were created with 15 ng of cfDNA.

### Preparation of patient and healthy honor cfDNA samples

Plasma processing and cfDNA extraction was performed according to the standard operating procedures at Personalis. Healthy donor plasma was isolated from whole blood collected in Streck Cell-Free DNA BCTs (Streck, La Vista, NE) and clarified using sequential centrifugation at 1,600 RCF and 16,000 RCF, respectively. Patient plasma from vendors was thawed and then clarified at 15,000 RCF prior to cfDNA extraction. For all plasma samples, cfDNA was extracted from up to 4.1 mL clarified plasma using a commercially available kit (QIAGEN, Germantown, MD) and quantified using Cell-free DNA ScreenTape Analysis (Agilent Technologies, Santa Clara, CA). For the clinical sample performance and specificity studies, up to 30 ng cfDNA was included in the preparation of pre-enrichment libraries. For the contrived sample functional characterization study, patient cfDNA was serially diluted with a healthy donor cfDNA sample, and then pre-enrichment libraries were created using 15 ng of cfDNA.

### NeXT Personal cfDNA Library Preparation, Target Enrichment and Sequencing

Library preparation, target enrichment and sequencing of the cfDNA samples was performed according to the standard operating procedures at Personalis. Briefly, pre-enrichment libraries were prepared from 2 ng to 30 ng cfDNA input using a commercially available KAPA Kit (Roche Sequencing Solutions, Pleasanton, CA) and Personalis-optimized workflows. A Lunatic spectrophotometer (Unchained Labs, Pleasanton, CA) was used to quantify the pre-enrichment libraries. Up to 1500 ng of library DNA was enriched with NeXT Personal custom probe panels using proprietary modifications to a commercially available Twist kit and hybridization-capture workflow (Twist Bioscience, South San Francisco, CA). The post-enrichment libraries were significantly over-sequenced on NovaSeq instruments (Illumina, San Diego, CA) in order to optimize the number of unique molecules observed.

### NeXT Personal MRD Analysis

The results and data presented in this study utilize the production version of the NeXT Personal analysis pipelines, in use for the LDT version of the assay at the time of publication. Briefly, sequencing data from cfDNA libraries enriched with patient specific panels is used to form consensus reads of the captured molecules across the targeted regions. Proprietary grouping and filtering approaches ensure the formation of accurate read groupings prior to molecular consensus formation. A statistical analysis is undertaken to vet and aggregate the ctDNA signal across all MRD targets. A ctDNA positive call is made only when the p-value assigned by that statistical analysis is less than 0.001 (indicating a false positive rate of less than 1 in 1,000). The ctDNA signal is reported in PPM, calculated as the number of consensus molecules containing a tumor derived variant divided by the total number of consensus molecules covering MRD targets.

### Specificity Evaluation Using Simulated Panels

The specificity of the NeXT Personal platform was empirically examined using the 118 patient-specific panels described in the Clinical Sample Performance study (see results). Two complementary analyses were performed *in silico* with 23,600 simulated panels each. First, the simulated panels were generated according to the approach of Abbosh et al.2023. In brief, *in silico* panels were created via replacement of the original MRD variants in the 118 patient-specific panels, maintaining the size and trinucleotide error context of each panel. MRD targets were replaced by random sampling with bases within 50 bp of the actual target with similar coverage, passing NeXT Personal target selection requirements, and not indicated as known germline variants in dbSNP (build 146, https://www.ncbi.nlm.nih.gov/pmc/articles/PMC102496/). Sequencing data from the Specificity study was then processed through the standard NeXT Personal pipeline to detect any false positive calls. Second, the simulated panels were generated by re-shuffling the targets used across all panels in the Specificity study into new combinations that maintain the size and trinucleotide error context of each panel. Sequencing data for the new combinations of targets from the Specificity study was then re-processed through the standard NeXT Personal pipeline to detect any false positive calls.

### Statistical analysis

The studies performed in this validation were planned in consultation with and analyzed by an external statistical consultant (Stat4Ward, Pittsburgh, PA). Analysis was performed in R Statistical Software. All statistical tests were 2-sided unless otherwise stated. Detailed descriptions of the statistical analyses can be found in the supplemental material.

## Data Availability

All data produced in the present work are contained in the manuscript.

## ACKNOWLEDGEMENTS

The authors would like to thank Shuguang Huang, Zhengbo Wu, Joanne Yu and Jia Zhang at Stat4ward for their help with the study design and statistical analysis of the results. The authors would like to thank Sylvia Signorelli for helping to coordinate experiment logistics and sample preparation, as well as providing SOP oversight. The authors would like to thank Upasana Dutta, Mukul Nerlekar, Liberty Onia and Alex Winans for assisting with sample preparation, labeling and aliquoting. The authors would like to thank members of the operations team: Don Brown for helping with resources and timelines, Jennifer Claros and team for histology support, Ayupe Obad, Barbra Altawil, Casey Fiel, Kaleigh Thorne and the NeXT Personal RUO team for assisting with sample processing, and the sequencing team for loading the instruments. The authors would like to thank Misha Agarwal, Stephen Fairclough, Kate Gerhardt, Brandon Giang, Vijay Gillella, Aniruddhan Govindaraman, Edward Gow, Peter Morrison, Shilpa Nalla, Karthikeyan Swaminathan, Tam Truong and Chunwei Wang, for informatics and software support. The authors would like to thank Manju Chinnappa and Galina Zybina for providing feedback on the study plan, as well as Charles Abbott, Sean Boyle and Erich Jaeger for their help reviewing the manuscript. The authors would also like to thank John West for his contributions to the concept of the NeXT Personal assay.

## AUTHOR CONTRIBUTIONS

GB, NU, JSS, RB, YC, DN, JML, participated in the study design creation. JN, GB, NU, JSS, EA, JA, RB, LJG, RC, JML, participated in the study design review. JN, JML, participated in the validation plan creation. GB, MH, QZ, participated in the validation plan review. JN, JLa, TL, CM, RS, JML, participated in the validation execution oversight. TL, RC, RV, participated in the validation experiment execution. JN, MH, SM, JML, participated in the pre-validation studies planning. MH, SM, participated in the pre-validation studies execution. JN, QZ, SM, TXC, JML, participated in the supporting studies planning. QZ, SM, TXC, participated in the supporting studies execution. JH, JLi, TC, participated in the informatic pipeline support. JN, GB, JH, CL, FCPN, RMP, MH, QZ, TC, ROC, JML, participated in the data analysis and interpretation. GB, JML, participated in the statistical analysis. JN, GB, JH, CL, ROC, JML, participated in the manuscript writing and revisions. FCPN, RMP, JLa, NU, EA, YC, DN, participated in the manuscript review.

## SUPPLEMENTAL MATERIALS

### Supplemental Results

**Supplemental Figure S1:**
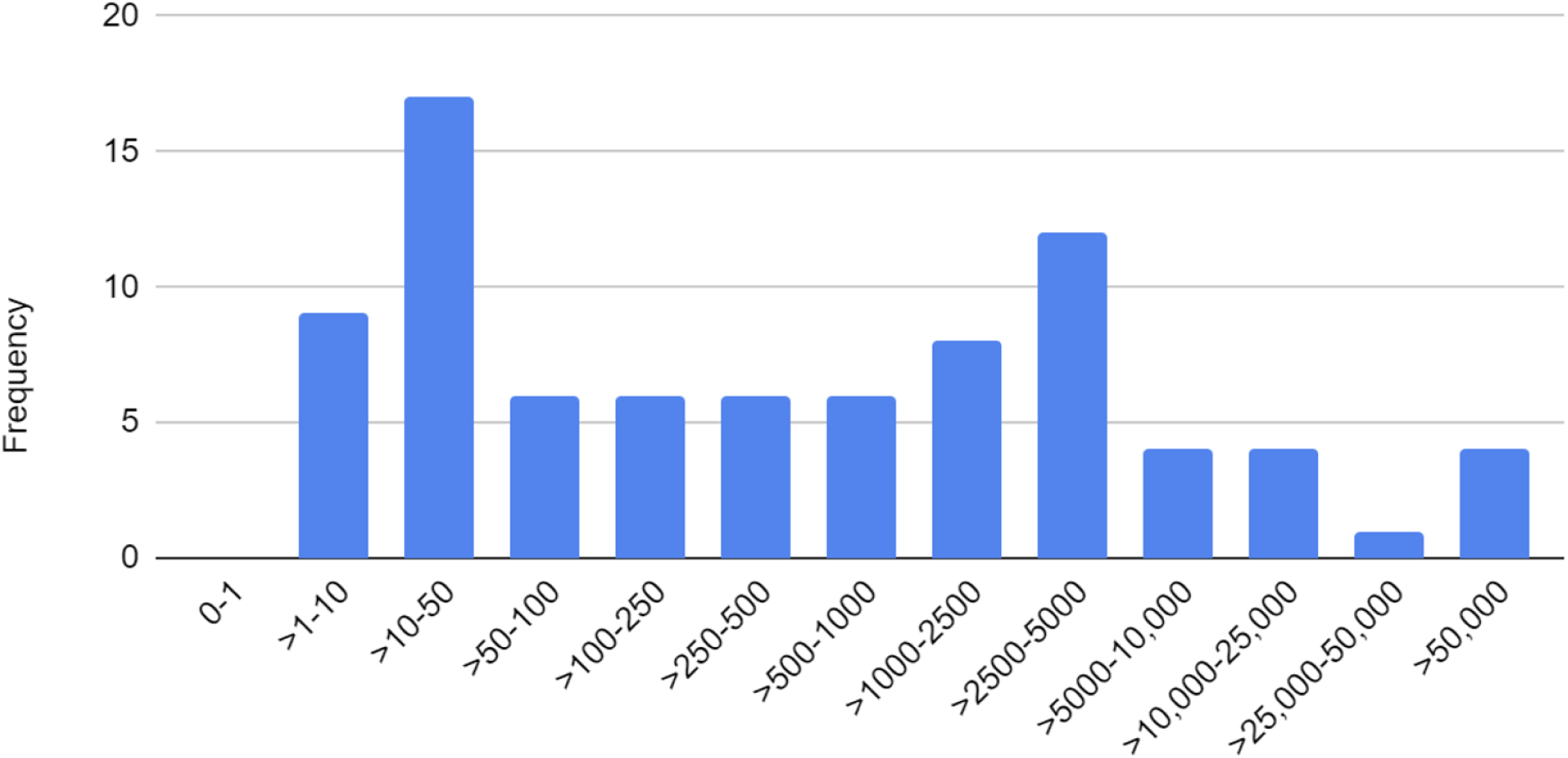
Histogram of the ctDNA signals (PPM) for clinical samples with NeXT Personal positive ctDNA calls. Levels of 25, 1,000, and 25,000 PPM were selected for the precision study as they are representative of low, medium, and high positive signals observed in clinical samples.

#### Interfering Substances: Hemoglobin

Another common contaminant in the plasma fraction is hemoglobin, released by the hemolysis of red blood cells. To study the effect of hemoglobin on the NeXT Personal assay, hemoglobin was added into normal blood from three donors at 0, 0.5, 1, 2, and 4 mg/mL. The highest amount (4 mg/mL) corresponds to nearly 3% hemolysis, and is 4 times as high as the allowable limit for laboratory processing [46]. After hemoglobin addition, the blood samples were processed according to the standard operating procedures for the NeXT Personal assay.

Measurements of hemoglobin before and after cfDNA extraction showed that hemoglobin was largely removed during extraction, resulting in no effect on cfDNA extraction or library yields, even at the highest amount of hemoglobin addition (Figure S2). We conclude that even with the presence of substantial free hemoglobin present in the patient blood donor sample, the NeXT Personal assay is not materially affected.

**Supplemental Figure S2:**
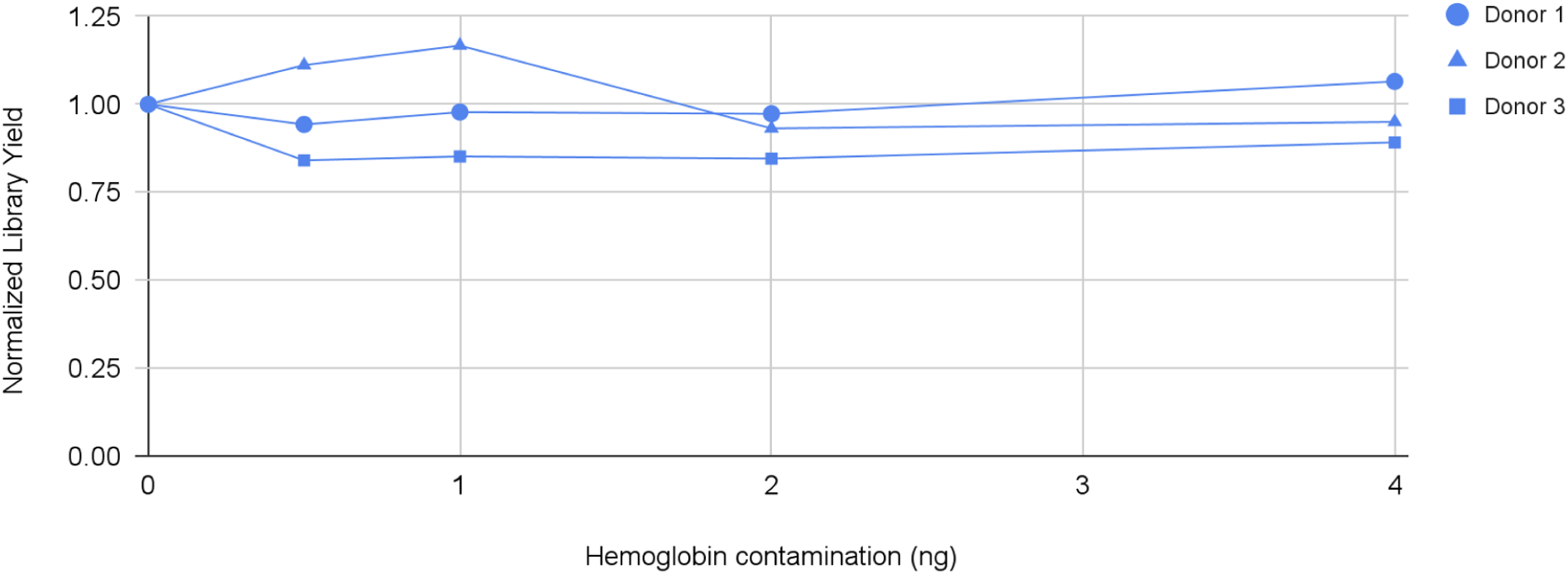
Impact of hemoglobin contamination on cfDNA library yield.

#### Interfering Substances: Nucleic acid extraction wash buffer

A risk-based assessment of the overall NeXT Personal lab process identified contamination of extracted cfDNA by the wash buffer from the step prior to elution as the highest potential risk for interfering substances resulting from the protocol. To demonstrate the robustness of the assay to this potential interferent, we added this wash buffer into purified cfDNA at a range of concentrations, spanning and exceeding those considered likely to occur. As shown in Figure S3, there was no impact on cfDNA library yield due to the presence of this potential interferent. Sequencing of the libraries showed no significant changes in error rates in the presence of the interferent (data not shown).

**Supplemental Figure S3.**
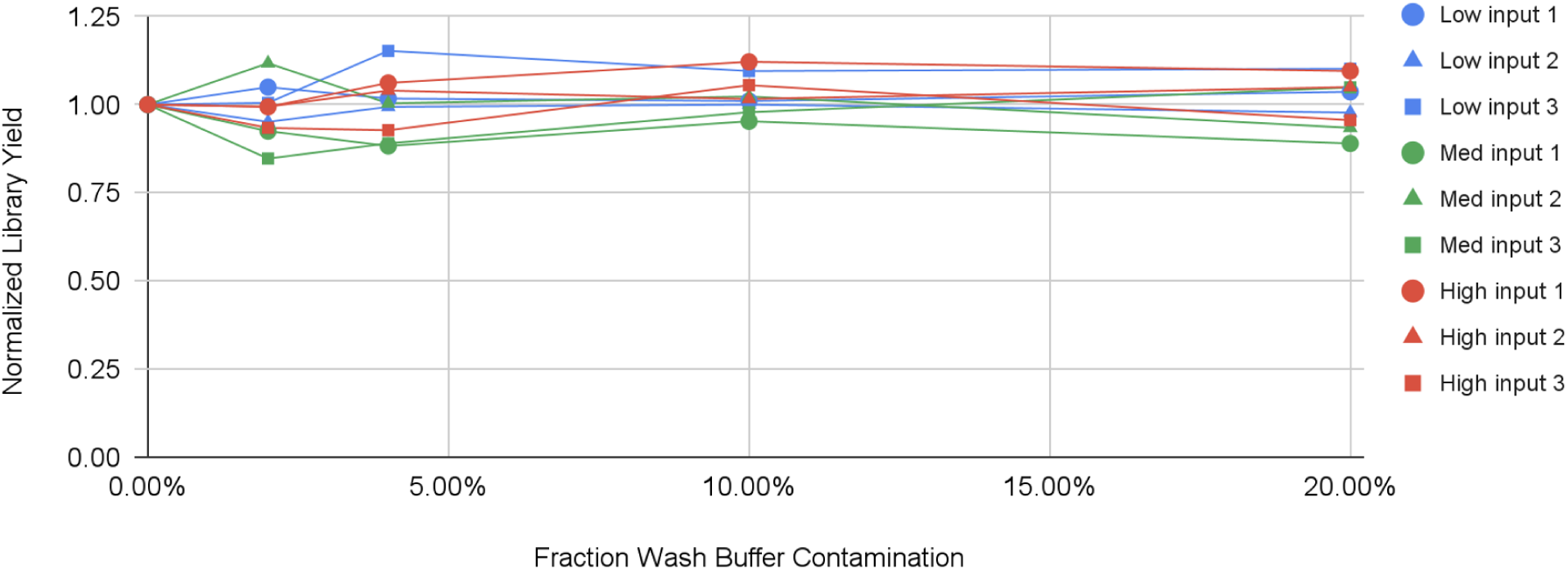
Effects of nucleic acid extraction wash buffer contamination on library yield.

**Supplemental Table S1.**
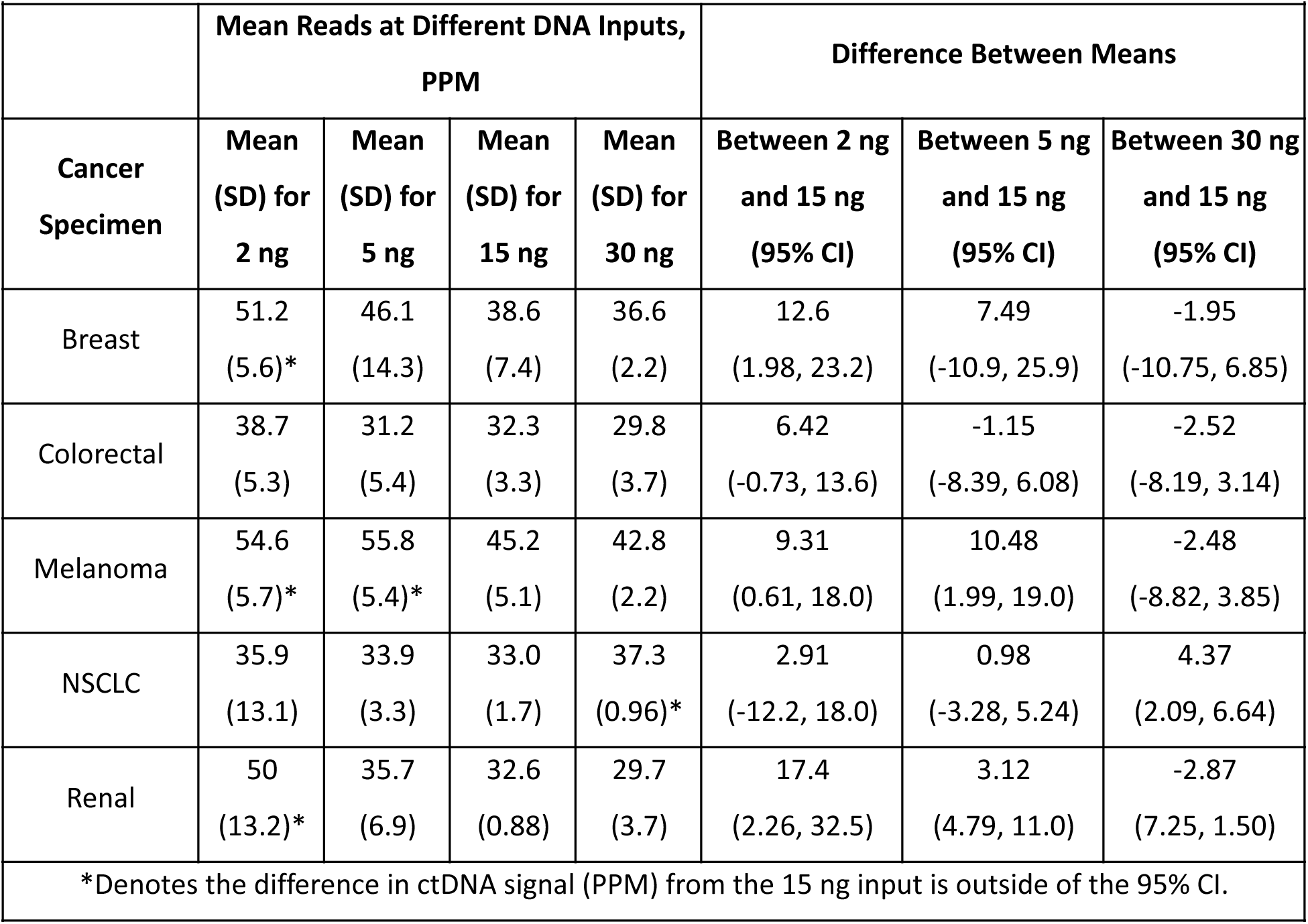
ctDNA measurements at various cfDNA input amounts, compared to 15 ng.

For the melanoma sample, the mean ctDNA signal at 5 ng input was significantly higher than at 15 ng input, but this difference was within the 25% TAE of our analytical range. In the NSCLC sample, the mean ctDNA signal at 30 ng input was also significantly higher than at 15 ng input, but we attribute this to the atypically small standard deviation (CV=2.6%) associated with those replicates. The actual deviation, 13.0%, is well within the 25% allowable error.

### Supplemental Methods

#### Accuracy Calculation

For the accuracy study, the reported confidence interval (CI) is the 95% CI, which is equivalent to 1.956 standard deviations above and below the mean. The assay sensitivity was calculated as the number of true-positive results divided by the sum of true-positive and false-negative results; assay specificity was calculated as the number of true-negative results divided by the sum of true-negatives and false-positive results; the positive predictive value was calculated as the number of true-positive results divided by the sum of true-positive and false-positive results; and the negative predictive value was calculated as the number of true-negative results divided by the sum of false-negative plus true-negative results.

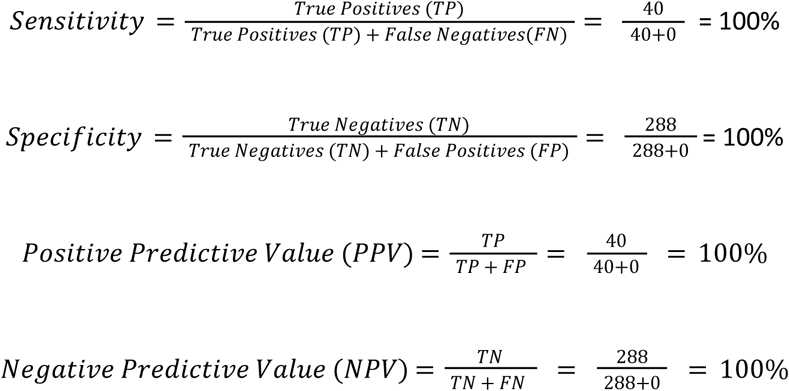

#### Limit of Detection Calculation

The limit of detection (LOD_95_) is defined by convention as the ctDNA concentration at which 95% of measurement results give a positive outcome [44]. The LOD_95_ is reported in units of parts per million (PPM).

In this LOD study, the contrived sample system was used to create a series of 7 low positive samples with known tumor mutation concentrations ranging from 0.8 PPM to 8.2 PPM. Each sample was run 5 times by 2 operators, for a total of 70 runs. The 5 replicates were enriched by the operator over 2 days, using different lots of the hybridization-capture enrichment kit.

The results for each reagent lot were plotted as the standard deviation (SD) of ctDNA signal levels versus the measured ctDNA signal. Using the precision profile approach [44], a third order polynomial equation was fit to each plot, and the best fit coefficients *B*_1_, *B*_2_, *B*_3_, and *B*_4_ were obtained (Eq. 1).

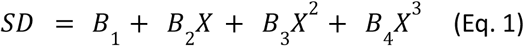

where the polynomial coefficients were obtained as shown in Table S2.

**Table S2.** Polynomial coefficients for reagent lots A and B.

| Table S2. Polynomial coefficients for reagent lots A and B |  |  |
| --- | --- | --- |
| Coefficient | Reagent Lot A | Reagent Lot B |
| B1 | -0.6726 | 0.7324 |
| B2 | 1.2806 | -0.3672 |
| B3 | -0.2190 | 0.2099 |
| B4 | 0.01249 | -0.0198 |

Using Eq. 2, a trial LOD was calculated for mean detection threshold and then numerically iterated to the new value until the result converged.

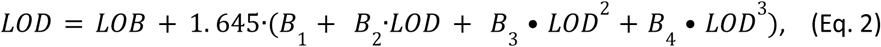

In Eq. 2, 1.645 represents the shift that would put 95% of the normal distribution of signals above the LOB (0.72 PPM per above) when the SD=1. The actual SD can be found from Eq.1 using the LOD for X. Since the highest LOD for the reagent lots is chosen for the final LOD, the final LOD is 3.45 PPM.

**Table S3.** Calculated LOD for Each Reagent Lot.

| Table S3. Calculated LOD for Each Reagent Lot |  |  |
| --- | --- | --- |
|  | Reagent Lot A | Reagent Lot B |
| LOD (PPM) | 3.45 | 1.80 |

## REFERENCES

1. Ococks E, Frankell AM, Masque Soler N, Grehan N, Northrop A, Coles H, Redmond AM, Devonshire G, Weaver JMJ, Hughes C, Lehovsky K, Blasko A, Nutzinger B, et al. Longitudinal tracking of 97 esophageal adenocarcinomas using liquid biopsy sampling. Ann Oncol. 2021; 32: 522–32. doi: 10.1016/j.annonc.2020.12.010.

2. Wan JCM, Mughal TI, Razavi P, Dawson SJ, Moss EL, Govindan R, Tan IB, Yap YS, Robinson WA, Morris CD, Besse B, Bardelli A, Tie J, et al. Liquid biopsies for residual disease and recurrence. Med. 2021; 2: 1292–313. doi: 10.1016/j.medj.2021.11.001.

3. Chaudhuri AA, Chabon JJ, Lovejoy AF, Newman AM, Stehr H, Azad TD, Khodadoust MS, Esfahani MS, Liu CL, Zhou L, Scherer F, Kurtz DM, Say C, et al. Early Detection of Molecular Residual Disease in Localized Lung Cancer by Circulating Tumor DNA Profiling. Cancer Discov. 2017; 7: 1394–403. doi: 10.1158/2159-8290.CD-17-0716.

4. Larribere L, Martens UM. Advantages and Challenges of Using ctDNA NGS to Assess the Presence of Minimal Residual Disease (MRD) in Solid Tumors. Cancers (Basel). 2021; 13. doi: 10.3390/cancers13225698.

5. Coombes RC, Page K, Salari R, Hastings RK, Armstrong A, Ahmed S, Ali S, Cleator S, Kenny L, Stebbing J, Rutherford M, Sethi H, Boydell A, et al. Personalized Detection of Circulating Tumor DNA Antedates Breast Cancer Metastatic Recurrence. Clin Cancer Res. 2019; 25: 4255–63. doi: 10.1158/1078-0432.CCR-18-3663.

6. Sausen M, Phallen J, Adleff V, Jones S, Leary RJ, Barrett MT, Anagnostou V, Parpart-Li S, Murphy D, Kay Li Q, Hruban CA, Scharpf R, White JR, et al. Clinical implications of genomic alterations in the tumour and circulation of pancreatic cancer patients. Nat Commun. 2015; 6: 7686. doi: 10.1038/ncomms8686.

7. Abbosh C, Birkbak NJ, Wilson GA, Jamal-Hanjani M, Constantin T, Salari R, Le Quesne J, Moore DA, Veeriah S, Rosenthal R, Marafioti T, Kirkizlar E, Watkins TBK, et al. Phylogenetic ctDNA analysis depicts early-stage lung cancer evolution. Nature. 2017; 545: 446–51. doi: 10.1038/nature22364.

8. Dyrskjt L, Laliotis Md PG, Nordentoft I, Birkenkamp-Demtřder K, Viborg Lindskrog S, Lamy P, White E, Pajak N, Andreasen TG, Dutta P, Sharma S, Calhoun M, ElNaggar A, et al. Utility of ctDNA in predicting outcome and pathological complete response in patients with bladder cancer as a guide for selective bladder preservation strategies. Journal of Clinical Oncology. 2023; 41: 563-. doi: 10.1200/JCO.2023.41.6_suppl.563.

9. Gale D, Heider K, Ruiz-Valdepenas A, Hackinger S, Perry M, Marsico G, Rundell V, Wulff J, Sharma G, Knock H, Castedo J, Cooper W, Zhao H, et al. Residual ctDNA after treatment predicts early relapse in patients with early-stage non-small cell lung cancer. Ann Oncol. 2022; 33: 500–10. doi: 10.1016/j.annonc.2022.02.007.

10. Tie J, Wang Y, Cohen J, Li L, Hong W, Christie M, Wong HL, Kosmider S, Wong R, Thomson B, Choi J, Fox A, Field K, et al. Circulating tumor DNA dynamics and recurrence risk in patients undergoing curative intent resection of colorectal cancer liver metastases: A prospective cohort study. PLoS Med. 2021; 18: e1003620. doi: 10.1371/journal.pmed.1003620.

11. Tan L, Sandhu S, Lee RJ, Li J, Callahan J, Ftouni S, Dhomen N, Middlehurst P, Wallace A, Raleigh J, Hatzimihalis A, Henderson MA, Shackleton M, et al. Prediction and monitoring of relapse in stage III melanoma using circulating tumor DNA. Ann Oncol. 2019; 30: 804–14. doi: 10.1093/annonc/mdz048.

12. Tie J, Wang Y, Tomasetti C, Li L, Springer S, Kinde I, Silliman N, Tacey M, Wong HL, Christie M, Kosmider S, Skinner I, Wong R, et al. Circulating tumor DNA analysis detects minimal residual disease and predicts recurrence in patients with stage II colon cancer. Sci Transl Med. 2016; 8: 346ra92. doi: 10.1126/scitranslmed.aaf6219.

13. Garcia-Murillas I, Chopra N, Comino-Mendez I, Beaney M, Tovey H, Cutts RJ, Swift C, Kriplani D, Afentakis M, Hrebien S, Walsh-Crestani G, Barry P, Johnston SRD, et al. Assessment of Molecular Relapse Detection in Early-Stage Breast Cancer. JAMA Oncol. 2019; 5: 1473–8. doi: 10.1001/jamaoncol.2019.1838.

14. Pereira E, Camacho-Vanegas O, Anand S, Sebra R, Catalina Camacho S, Garnar-Wortzel L, Nair N, Moshier E, Wooten M, Uzilov A, Chen R, Prasad-Hayes M, Zakashansky K, et al. Personalized Circulating Tumor DNA Biomarkers Dynamically Predict Treatment Response and Survival In Gynecologic Cancers. PLoS One. 2015; 10: e0145754. doi: 10.1371/journal.pone.0145754.

15. Azad TD, Chaudhuri AA, Fang P, Qiao Y, Esfahani MS, Chabon JJ, Hamilton EG, Yang YD, Lovejoy A, Newman AM, Kurtz DM, Jin M, Schroers-Martin J, et al. Circulating Tumor DNA Analysis for Detection of Minimal Residual Disease After Chemoradiotherapy for Localized Esophageal Cancer. Gastroenterology. 2020; 158: 494–505 e6. doi: 10.1053/j.gastro.2019.10.039.

16. Sato S, Nakamura Y, Oki E, Yoshino T. Molecular Residual Disease-guided Adjuvant Treatment in Resected Colorectal Cancer: Focus on CIRCULATE-Japan. Clin Colorectal Cancer. 2023; 22: 53–8. doi: 10.1016/j.clcc.2022.12.001.

17. <sanz-garcia modifying therapy w ctdna 2022.pdf>. doi:

18. Naidoo M, Gibbs P, Tie J. ctDNA and Adjuvant Therapy for Colorectal Cancer: Time to Re-Invent Our Treatment Paradigm. Cancers (Basel). 2021; 13. doi: 10.3390/cancers13020346.

19. Tie J, Cohen JD, Lahouel K, Lo SN, Wang Y, Kosmider S, Wong R, Shapiro J, Lee M, Harris S, Khattak A, Burge M, Harris M, et al. Circulating Tumor DNA Analysis Guiding Adjuvant Therapy in Stage II Colon Cancer. N Engl J Med. 2022; 386: 2261–72. doi: 10.1056/NEJMoa2200075.

20. Almodovar K, Iams WT, Meador CB, Zhao Z, York S, Horn L, Yan Y, Hernandez J, Chen H, Shyr Y, Lim LP, Raymond CK, Lovly CM. Longitudinal Cell-Free DNA Analysis in Patients with Small Cell Lung Cancer Reveals Dynamic Insights into Treatment Efficacy and Disease Relapse. J Thorac Oncol. 2018; 13: 112–23. doi: 10.1016/j.jtho.2017.09.1951.

21. Ulrich B, Pradines A, Mazieres J, Guibert N. Detection of Tumor Recurrence via Circulating Tumor DNA Profiling in Patients with Localized Lung Cancer: Clinical Considerations and Challenges. Cancers (Basel). 2021; 13. doi: 10.3390/cancers13153759.

22. Kasi PM, Fehringer G, Taniguchi H, Starling N, Nakamura Y, Kotani D, Powles T, Li BT, Pusztai L, Aushev VN, Kalashnikova E, Sharma S, Malhotra M, et al. Impact of Circulating Tumor DNA-Based Detection of Molecular Residual Disease on the Conduct and Design of Clinical Trials for Solid Tumors. JCO Precis Oncol. 2022; 6: e2100181. doi: 10.1200/PO.21.00181.

23. Crupi E, de Padua TC, Marandino L, Raggi D, Dyrskjot L, Spiess PE, Sonpavde GP, Kamat AM, Necchi A. Circulating tumor DNA as a Predictive and Prognostic Biomarker in the Perioperative Treatment of Muscle-invasive Bladder Cancer: A Systematic Review. Eur Urol Oncol. 2023. doi: 10.1016/j.euo.2023.05.012.

24. Bratman SV, Yang SYC, Iafolla MAJ, Liu Z, Hansen AR, Bedard PL, Lheureux S, Spreafico A, Razak AA, Shchegrova S, Louie M, Billings P, Zimmermann B, et al. Personalized circulating tumor DNA analysis as a predictive biomarker in solid tumor patients treated with pembrolizumab. Nat Cancer. 2020; 1: 873–81. doi: 10.1038/s43018-020-0096-5.

25. Assaf ZJF, Zou W, Fine AD, Socinski MA, Young A, Lipson D, Freidin JF, Kennedy M, Polisecki E, Nishio M, Fabrizio D, Oxnard GR, Cummings C, et al. A longitudinal circulating tumor DNA-based model associated with survival in metastatic non-small-cell lung cancer. Nat Med. 2023; 29: 859–68. doi: 10.1038/s41591-023-02226-6.

26. Lipsyc-Sharf M, de Bruin EC, Santos K, McEwen R, Stetson D, Patel A, Kirkner GJ, Hughes ME, Tolaney SM, Partridge AH, Krop IE, Knape C, Feger U, et al. Circulating Tumor DNA and Late Recurrence in High-Risk Hormone Receptor-Positive, Human Epidermal Growth Factor Receptor 2-Negative Breast Cancer. J Clin Oncol. 2022; 40: 2408–19. doi: 10.1200/JCO.22.00908.

27. Araujo DV, Bratman SV, Siu LL. Designing circulating tumor DNA-based interventional clinical trials in oncology. Genome Med. 2019; 11: 22. doi: 10.1186/s13073-019-0634-x.

28. US Food and Drug Administration. Use of Circulating Tumor DNA for Early-Stage Solid Tumor Drug Development - Guidance for Industry. 2022. https://www.fda.gov/regulatory-information/search-fda-guidance-documents/use-circulating-tumor-deoxyribonucleic-acid-early-stage-solid-tumor-drug-development-draft-guidance. Accessed Dec. 7, 2023.

29. Parikh AR, Mojtahed A, Schneider JL, Kanter K, Van Seventer EE, Fetter IJ, Thabet A, Fish MG, Teshome B, Fosbenner K, Nadres B, Shahzade HA, Allen JN, et al. Serial ctDNA Monitoring to Predict Response to Systemic Therapy in Metastatic Gastrointestinal Cancers. Clin Cancer Res. 2020; 26: 1877–85. doi: 10.1158/1078-0432.CCR-19-3467.

30. Tie J, Kinde I, Wang Y, Wong HL, Roebert J, Christie M, Tacey M, Wong R, Singh M, Karapetis CS, Desai J, Tran B, Strausberg RL, et al. Circulating tumor DNA as an early marker of therapeutic response in patients with metastatic colorectal cancer. Ann Oncol. 2015; 26: 1715–22. doi: 10.1093/annonc/mdv177.

31. Parkinson CA, Gale D, Piskorz AM, Biggs H, Hodgkin C, Addley H, Freeman S, Moyle P, Sala E, Sayal K, Hosking K, Gounaris I, Jimenez-Linan M, et al. Exploratory Analysis of TP53 Mutations in Circulating Tumour DNA as Biomarkers of Treatment Response for Patients with Relapsed High-Grade Serous Ovarian Carcinoma: A Retrospective Study. PLoS Med. 2016; 13: e1002198. doi: 10.1371/journal.pmed.1002198.

32. Wei T, Zhang Q, Li X, Su W, Li G, Ma T, Gao S, Lou J, Que R, Zheng L, Bai X, Liang T. Monitoring Tumor Burden in Response to FOLFIRINOX Chemotherapy Via Profiling Circulating Cell-Free DNA in Pancreatic Cancer. Mol Cancer Ther. 2019; 18: 196–203. doi: 10.1158/1535-7163.MCT-17-1298.

33. Chehrazi-Raffle A, Muddasani R, Dizman N, Hsu J, Meza L, Zengin ZB, Malhotra J, Chawla N, Dorff T, Contente-Cuomo T, Dinwiddie D, McDonald BR, McDaniel T, et al. Ultrasensitive Circulating Tumor DNA Pilot Study Distinguishes Complete Response and Partial Response With Immunotherapy in Patients With Metastatic Renal Cell Carcinoma. JCO Precis Oncol. 2023; 7: e2200543. doi: 10.1200/PO.22.00543.

34. Velimirovic M, Juric D, Niemierko A, Spring L, Vidula N, Wander SA, Medford A, Parikh A, Malvarosa G, Yuen M, Corcoran R, Moy B, Isakoff SJ, et al. Rising Circulating Tumor DNA As a Molecular Biomarker of Early Disease Progression in Metastatic Breast Cancer. JCO Precis Oncol. 2020; 4: 1246–62. doi: 10.1200/PO.20.00117.

35. Chan HT, Nagayama S, Otaki M, Chin YM, Fukunaga Y, Ueno M, Nakamura Y, Low SK. Tumor-informed or tumor-agnostic circulating tumor DNA as a biomarker for risk of recurrence in resected colorectal cancer patients. Front Oncol. 2022; 12: 1055968. doi: 10.3389/fonc.2022.1055968.

36. Finkle JD, Boulos H, Driessen TM, Lo C, Blidner RA, Hafez A, Khan AA, Lozac’hmeur A, McKinnon KE, Perera J, Zhu W, Dowlati A, White KP, et al. Validation of a liquid biopsy assay with molecular and clinical profiling of circulating tumor DNA. NPJ Precis Oncol. 2021; 5: 63. doi: 10.1038/s41698-021-00202-2.

37. Woodhouse R, Li M, Hughes J, Delfosse D, Skoletsky J, Ma P, Meng W, Dewal N, Milbury C, Clark T, Donahue A, Stover D, Kennedy M, et al. Clinical and analytical validation of FoundationOne Liquid CDx, a novel 324-Gene cfDNA-based comprehensive genomic profiling assay for cancers of solid tumor origin. PLoS One. 2020; 15: e0237802. doi: 10.1371/journal.pone.0237802.

38. Zhao J, Reuther J, Scozzaro K, Hawley M, Metzger E, Emery M, Chen I, Barbosa M, Johnson L, O’Connor A, Washburn M, Hartje L, Reckase E, et al. Personalized Cancer Monitoring Assay for the Detection of ctDNA in Patients with Solid Tumors. Mol Diagn Ther. 2023; 27: 753–68. doi: 10.1007/s40291-023-00670-1.

39. Keller L, Belloum Y, Wikman H, Pantel K. Clinical relevance of blood-based ctDNA analysis: mutation detection and beyond. Br J Cancer. 2021; 124: 345–58. doi: 10.1038/s41416-020-01047-5.

40. Parsons HA, Rhoades J, Reed SC, Gydush G, Ram P, Exman P, Xiong K, Lo CC, Li T, Fleharty M, Kirkner GJ, Rotem D, Cohen O, et al. Sensitive Detection of Minimal Residual Disease in Patients Treated for Early-Stage Breast Cancer. Clin Cancer Res. 2020; 26: 2556–64. doi: 10.1158/1078-0432.CCR-19-3005.

41. Moding EJ, Nabet BY, Alizadeh AA, Diehn M. Detecting Liquid Remnants of Solid Tumors: Circulating Tumor DNA Minimal Residual Disease. Cancer Discov. 2021; 11: 2968–86. doi: 10.1158/2159-8290.CD-21-0634.

42. Dang DK, Park BH. Circulating tumor DNA: current challenges for clinical utility. J Clin Invest. 2022; 132. doi: 10.1172/JCI154941.

43. Fakih M, Sandhu J, Wang C, Kim J, Chen YJ, Lai L, Melstrom K, Kaiser A. Evaluation of Comparative Surveillance Strategies of Circulating Tumor DNA, Imaging, and Carcinoembryonic Antigen Levels in Patients With Resected Colorectal Cancer. JAMA Netw Open. 2022; 5: e221093. doi: 10.1001/jamanetworkopen.2022.1093.

44. Clinical and Laboratory Standards Institute. Detection Capability for Clinical Laboratory Measurement Procedures; Approved Guideline—Second Edition 2012. 950 West Valley Road, Suite 2500 Wayne, PA 19087 USA.

45. Abbosh C, Frankell AM, Harrison T, Kisistok J, Garnett A, Johnson L, Veeriah S, Moreau M, Chesh A, Chaunzwa TL, Weiss J, Schroeder MR, Ward S, et al. Tracking early lung cancer metastatic dissemination in TRACERx using ctDNA. Nature. 2023; 616: 553–62. doi: 10.1038/s41586-023-05776-4.

46. Center for Disease Control and Prevention. Division of Vector-Borne Diseases A Quick-Reference Tool for Hemolysis Status. https://www.cdc.gov/ncezid/dvbd/specimensub/hemolysis-palette.html. Accessed Dec. 7, 2023.

